# Toxicities of PD-1/PD-L1 Inhibitors: A Systematic Review and Meta-Analysis of Randomized Controlled Trials

**DOI:** 10.1101/2020.07.16.20155309

**Authors:** Xiangyi Kong, Li Chen, Ryan J Sullivan, Zhihong Qi, Yulu Liu, Yi Fang, Lin Zhang, Jing Wang

**Author notes:** **Corresponding Authors Jing Wang**, Department of Breast Surgical Oncology, National Cancer Center/National Clinical Research Center for Cancer/Cancer Hospital, Chinese Academy of Medical Sciences and Peking Union Medical College, Beijing, 100021, China, **Lin Zhang**, School of Public Health, Chinese Academy of Medical Sciences and Peking Union Medical College, Beijing, 100021, China; Melbourne School of Population and Global Health, The University of Melbourne, Victoria, 3010, Australia; Centre of Cancer Research, Victorian Comprehensive Cancer Centre, Melbourne, Victoria, 3010, Australia, **Yi Fang**, Department of Breast Surgical Oncology, National Cancer Center/National Clinical Research Center for Cancer/Cancer Hospital, Chinese Academy of Medical Sciences and Peking Union Medical College, Beijing, 100021, China. **These 2 authors contributed equally to this article.** (Xiangyi Kong, Li Chen).

## Abstract

**Background:** Immunotherapy, especially immune-checkpoint inhibitors (PD-1 and PD-L1 inhibitors), is now one of the mainstays of cancer treatment. Several studies have analyzed treatment-related toxicities of immunotherapy. However, small sample size, rough and unspecific stratification, and lack of comparison (pure sing-arm studies) are common limitations. Detailed organ- and system-specific toxicities remain not clear enough.

**Methods:** We systematically searched PubMed, Embase, Cochrane Library, Web of Science and others (CNKI), from database inception to Mar 31, 2020, for randomized controlled trials (RCTs) related to PD-1/PD-L1 inhibitors that had available toxicity data. We excluded non-randomized trials. The primary endpoint was to assess the difference in the incidences of toxicities between cancer patients who did and did not receive PD-1/PD-L1 inhibitors. We calculated the pooled relative risks (RRs) and corresponding 95% confidence intervals (95% CIs) using a random-effects model and assessed the heterogeneity between different groups. The subgroup analyses were conducted based on toxicity grade (severity), system and organ, treatment regimens in the intervention arm and control arm, PD-1/PD-L1 inhibitor drug type, and cancer histotype. We applied the five-point Jadad ranking system to evaluate the quality of the selected studies. We performed multivariate meta-regression analyses to explore the proportion of between-study variance.

**Findings:** A total of 29 eligible RCTs including 8067 patients were selected for the meta-analysis based on specified inclusion and exclusion criteria. Patients treated with PD-1/PD-L1 inhibitors were at lower risks of overall toxicities (all grades: RR 0.91, 95% CI 0.89-0.92; grade 3∼4: RR 0.76, 95% CI 0.74-0.78), including gastrointestinal toxicity (all grades: RR 0.68, 95% CI 0.60-0.77; grade 3∼4: RR 0.71, 95% CI 0.43-1.20), hematologic toxicity (all grades: RR 0.66, 95% CI 0.51-0.85; grade 3∼4: RR 0.55, 95% CI 0.37-0.83), and treatment event leading to discontinuation (all grades: RR 0.78, 95% CI 0.72-0.84; grade 3∼4: RR 0.58, 95% CI 0.49-0.67); but were at higher risks for respiratory toxicity (all grades: RR 1.74, 95% CI 1.33-2.28; grade 3∼4: RR 1.92, 95% CI 1.45-2.55) and endocrine toxicity (all grades: RR 1.70, 95% CI 0.62-4.69; grade 3∼4: RR 1.29, 95% CI 0.45-3.69). The subgroup analyses indicated that when compared with the control, toxicity comparison tendency for PD-1/PD-L1 inhibitors varied with the toxicity grade, affected system and organ, treatment regimens in the intervention arm and control arm, drug type, and cancer histotype. The male-female ratio was a statistically significant variable in the Meta-Regression analysis (*I*^2^=89.1, *τ*^2^=0.01, and P=0.001).

**Interpretation:** For most toxicity types based on system and organ, the incidence proportions for patients in the intervention arm were lower than those in the control arm, which suggested the general safety of PD-1/PD-L1 inhibitors against conventional chemotherapy and CTLA-4 inhibitors. However, for some specific toxicities including respiratory, cutaneous, and endocrine toxicities, the case was the opposite. The toxicity grade, system and organ, treatment regimens, drug type, and cancer histotype were all influencing factors. To our knowledge, this was by far the most comprehensive meta-analysis of RCTs on toxicities of immune-checkpoint inhibitors. Future research should focus on taking effective targeted measures to decrease the risks of different toxicities for different patient populations.

**Author Contributions:** Conceptualization, Xiangyi Kong, Yi Fang, Jing Wang; Data Collection, Xiangyi Kong, Li Chen; Methodology, Lin Zhang, Zhihong Qi, Yulu Liu; Supervision, Lin Zhang, Ryan J Sullivan; Writing - original draft, Xiangyi Kong, Lin Zhang; Writing - review & editing, Ryan J Sullivan, Xiangyi Kong, Lin Zhang. All authors have read and agreed to the published version of the manuscript.

**Funding:** This work was supported by the Beijing Municipal Natural Science Foundation (No. 7204293 and No. 19G10077), the Special Research Fund for Central Universities, Peking Union Medical College (No. 3332019053), the Beijing Hope Run Special Fund of Cancer Foundation of China (No. LC2019B03 and No. LC2019L07), the Natural Science Foundation of China (No. 81872160), and National Key R&D Program of China (No. 2018YFC1315000 and No. 2018YFC1315003). The funders have no conflicts of interests.

**Acknowledgments:** None.

**Conflicts of Interest:** The authors declare no conflict of interest.

**Ethical Approval:** The analysis followed the Strengthening the Reporting of Observational Studies in Epidemiology (STROBE) guidelines. National Cancer Center/National Clinical Research Center for Cancer/Cancer Hospital, Chinese Academy of Medical Sciences and Peking Union Medical College institutional review board approved this study as exempt.

**Registration:** We registered the research protocol with PROSPERO (registration number CRD42019135113).

**Copyright:** © 2020 The Author(s). Published by Elsevier Ltd. This is an Open Access article under the CC BY 4.0 license.

**Research in context**

Evidence before this study
PD-1/PD-L1 inhibitors have increasingly become a mainstay of cancer treatment in recent years. Growing evidence supports the existence of a different pattern of toxicities for PD-1/PD-L1 inhibitors in comparison with the conventional chemotherapy. However, the organ- and system-specific toxicity spectrum remained not clear enough. We hypothesized that the types and severities of such toxicities varied with the agents used as well as the cancer histotypes. We systematically searched PubMed, Embase, Cochrane Library, Web of Science and others (CNKI), from database inception to Mar 31, 2020, for randomized controlled trials (RCTs) related to PD-1/PD-L1 inhibitors that had available toxicity data. The search terms included “PD-1”, “programmed death receptor 1”, “PD-L1”, “programmed cell death 1 ligand 1”, “immunotherapy”, “immune checkpoint inhibitor”, “durvalumab”, “avelumab”, “ipilimumab”, “tremelimumab”, “nivolumab”, “pembrolizumab”, “atezolizumab”. A few previous meta-analyses investigated the incidences of immunotherapy-related toxicities among a variety of cancer histotypes, drugs, and dosing schedules. However, most of them were single-arm analyses, thus we could not conclude whether the toxicity incidence was higher or lower than patients who did not receive PD-1/PD-L1 inhibitors. Also, their stratification standards were easy and rough, hence not sufficient enough to provide a detailed toxicity spectrum.

Added value of this study
To the best of our knowledge, our study was by far the most comprehensive meta-analysis of randomized controlled trials (RCTs) on toxicities of PD-1 and PD-L1 inhibitors, providing evidence of a significant difference in the toxicities in PD-1/PD-L1 inhibitors and the control. Using published data from 29 RCTs, for more than 15162 patients with several types of advanced cancers, we showed that for most toxicity types, the incidence proportions for patients in the intervention arm were lower than those in the control arm, which suggested the general safety of PD-1/PD-L1 inhibitors. However, for some specific toxicities such as respiratory, cutaneous, and endocrine toxicities, the case was the opposite. The toxicities varied with the toxicity-grade, system and organ, treatment regimens, drug type, and cancer histotype.

Implications of all the available evidence
Our findings have several potential implications for clinical practice and future research. It is interesting that there are differences in the toxicity profiles in patients treated with PD-1/PD-L1 inhibitors based on specific drug types and cancer histotypes. Comparing with the control, when administering PD-1/PD-L1 inhibitors, possible respiratory toxicities (cough and dyspnea), mild cutaneous toxicities (pruritus, rash, and vitiligo), and endocrine toxicity (hypothyroidism and hyperthyroidism) should be given more attention. For instance, patients treated with pembrolizumab were more likely to experience rash, hepatitis, and pneumonitis than nivolumab; patients treated with PD-L1 inhibitors were at higher risks for developing cough and colitis. The first future research direction is to conduct a further meta-analysis to assess the heterogeneity of PD-1/PD-L1 inhibitor toxicities between different sexes, ages, races and Eastern Cooperative Oncology Group (ECOG) scores to obtain a better and unbiased estimation of the toxicities. The second direction should focus on taking effective targeted measures to decrease the risks of different toxicities for different patient populations.

## 1 Introduction

Monoclonal antibodies targeting immune checkpoints are able to restore antitumor immunity, thus reversing immune escape or evasion and promoting tumor cell death. Such antibodies include those targeting the cytotoxic T lymphocyte associated antigen 4 (CTLA-4)-CD28 and programmed cell death 1 (PD-1)–programmed cell death 1 ligand 1 (PD-L1) axes. The PD-1 pathway regulates the necessary balance between the stimulatory signals required for an effective immune response to external microorganisms and inhibitory signals for maintenance of self-tolerance [1]. This pathway also plays an important role in immune evasion from cancer-specific T cells [2]. The PD-1 ligands, PD-L1 and PD-L2, can be expressed on tumor cells or immune cells, including those infiltrating tumors. Activation of the PD-1/PD-L pathway leads to inhibition of the cytotoxic T cell response [3, 4]. Inhibiting the interaction of PD-1 and its ligands results in significant enhancement of T-cell function and therefore anti-tumor activity [5]. Anti-PD-1 antibodies such as nivolumab, pembrolizumab, and cemiplimab, as well as anti-PD-L1 antibodies such as avelumab, durvalumab and atezolizumab, have been developed. These anti-PD-1/PD-L1 agents have achieved great success over conventional treatments in many types of tumors and have been approved by the US FDA and the European Medicines Agency (EMA). Although anti-PD-1/PD-L1 have provoked a total paradigm shift in the treatment of oncological malignancies, a different pattern of toxicity and the emergence of a new spectrum of toxicities or toxicities have arisen in comparison with traditional chemotherapy agents of monoclonal antibodies. The frequency of toxicities is mainly dependent on the agents used but also on the specific characteristics of individual patients. The incidence of fatal cancer immunotherapy associated toxicities is estimated to be between 0.3% and 1.3% [6].

Toxicities can affect almost any organ, with varying frequencies and severities [7]. According to Martins et al.’s review [8], cutaneous toxicities affect between one-third and more than half of all patients receiving PD-1/PD-L1 inhibitors, among which, rash, pruritus and vitiligo are the most widely reported. Colitis is the most frequently observed toxicity in patients receiving ipilimumab, occurring in 10-20% of patients [9]. Pulmonary toxicities are often challenging to diagnose, especially among patients with lung cancer who also have pre-existing chronic lung disease. Anti-PD-1 or anti-PD-L1 antibody-induced pneumonitis is more frequent in the first-line setting and has both a greater incidence and severity in patients with NSCLC than in those with melanoma [10, 11]. Hypophysitis, a condition involving inflammation of the pituitary gland, is rare in patients receiving anti-PD-1 antibodies but much more common in those receiving ipilimumab, with an incidence of 12.0∼13.3% in the real-world setting [12, 13]. Nearly 20% of patients receiving anti-PD-1 antibodies present with thyroid dysfunction [14]. In a retrospective registry study of data from eight clinical centers [15], investigators estimated the prevalence of PD-1/PD-L1 inhibitors-induced myocarditis to be 1.14%; a toxicity associated with a high risk of death. Kao et al. published a retrospective cohort study describing the development of neurological complications with an incidence of 2.9% (10/347) in patients receiving anti-PD-1 antibodies [16]. Uveitis and sicca syndrome are the main ocular toxicities of PD-1/PD-L1 inhibitors reported in the literature [17]. In 2017, Cappelli et al. published a dedicated systematic review of the literature on rheumatological and musculoskeletal toxicities. These authors reported considerable variations in the reported incidences of arthralgia and myalgia, ranging from 1% to 43% and 1% to 20%, respectively [18]. Acute interstitial nephritis (AIN) is the most common renal toxicity, with an underlying pathogenesis that differs from that of other drug-related forms of AIN, in which a delayed hypersensitivity reaction is involved as opposed to a loss of self-tolerance, as is more commonly observed in patients receiving PD-1/PD-L1 inhibitors [19]. Other reported toxicities include encephalitis and/or aseptic meningitis, myasthenia gravis or necrotizing myositis, GBS, GBS-like syndromes and other inflammatory neuropathies, and hematologic toxicities.

Toxicities associated with immune checkpoint inhibitors are usually manageable, but they may sometimes lead to treatment withdrawal, and fulminant and fatal events can also occur [20]. A thorough understanding of toxicities, including the underlying pathogenesis, kinetics of appearance and clinical presentation, will not only help clinicians to manage these events more effectively but also enable assessments of the safety of treatment resumption after toxicity resolution [21]. Rare forms of toxicity are increasingly being reported in the medical literature, and clinicians must take into consideration the heterogeneous clinical presentations of patients with these events and the broad spectrum of affected organs. Since substantial variations exist in cancer histotype, drug and dosing schedule, and toxicity reporting criteria in the publication, ignoring these variations and missing data patterns in toxicity reporting can lead to inaccurate estimation of the true incidences of treatment-related toxicities associated with PD-1/PD-L1 inhibitors. As such, a series of meta-analyses have been conducted to try to provide a relatively stable conclusion (**Table S1**) [22-55]. They generally investigated the incidences of different treatment-related toxicities among a variety of cancer histotypes, drugs, and dosing schedules. However, three main limitations were nonnegligible for these meta-analyses: **1)** Most of them are single-arm analyses, thus we could not conclude whether the toxicity incidence is higher or lower than patients who did not receive PD-1/PD-L1 inhibitors; **2)** Although many of these studies considered stratification analysis by the spectrum of organs and systems affected by toxicities, the classification is not detailed and accurate; **3)** Among these studies, the most current literature search time is up to February 2019.

Considering the rapidly evolving clinical trial evidence and experience with PD-1/PD-L1 inhibitors over the past year and that many new clinical trial results have been published, an update is worthy. We herein performed a systematic review and meta-analysis of toxicities of the Food and Drug Administration (FDA)-approved PD-1/PD-L1 inhibitors in published clinical trials globally. We investigated the incidences of different treatment-related toxicities associated with these drugs, and we quantified the potential differences in toxicity incidences among a variety of toxicities and corresponding organs and systems affected, cancers, intervention regimens and drugs. We only included randomized controlled trials.

## 2 Methods

We followed PRISMA guidelines for this systematic review and meta-analysis [56]. We registered the research protocol with PROSPERO (registration number CRD42019135113). Two reviewers (K.X. and C.L.) independently undertook the literature search, assessment for eligibility, data extraction and qualitative assessment. Any inconsistencies between the two reviewers were reviewed by a third reviewer (L.Z.) and resolved by consensus.

### 2.1 Data Sources and Searches

A comprehensive literature search was conducted to identify all relevant articles. The databases interrogated were PubMed, Embase, Cochrane Library, Web of Science and others (CNKI). The dates searched were from the inception of each database Mar 31, 2020. Abstracts and presentations were also reviewed from all major conference proceedings, including American Society of Clinical Oncology (ASCO) and European Society for Medical Oncology (ESMO) from Jan 2010 to March 2020. Two investigators (K.X. and C.L.) independently searched the databases. The search terms included the following keywords: “PD-1”, “programmed death receptor 1”, “PD-L1”, “programmed cell death 1 ligand 1”, “immunotherapy”, “immune checkpoint inhibitor”, “durvalumab”, “avelumab”, “ipilimumab”, “tremelimumab”, “nivolumab”, “pembrolizumab”, “atezolizumab”.

We used the Cochrane highly sensitive search strategies for identifying randomized trials in PubMed and Embase [57].

Search filters used in PubMed (Sensitivity-maximizing version, 2008 revision):

#1 randomized controlled trial [pt]

#2 controlled clinical trial [pt]

#3 randomized [tiab]

#4 placebo [tiab]

#5 drug therapy [sh]

#6 randomly [tiab]

#7 trial [tiab]

#8 groups [tiab]

#9 #1 OR #2 OR #3 OR #4 OR #5 OR #6 OR #7 OR #8

#10 animals [mh] NOT humans [mh]

#11 #9 NOT #10

Search filters used in Embase:

‘crossover procedure’:de OR ‘double-blind procedure’:de OR ‘randomized controlled trial’:de OR ‘single-blind procedure’:de OR (random* OR factorial* OR crossover* OR cross NEXT/1 over* OR placebo* OR doubl* NEAR/1 blind* OR singl* NEAR/1 blind* OR assign* OR allocat* OR volunteer*):de,ab,ti

The search was extended by review of references of articles included in the final selection. We reviewed each publication and only the most recent and complete report of clinical trials was included. We combined the search results in a bibliographic management tool (EndNote) and used the Bramer method to eliminate duplicates.

### 2.2 Study Selection and Data Extraction

We identified all phase II and III randomized clinical trials (RCTs) with PD-1/PD-L1 inhibitors administered alone or in combination with chemotherapy, compared to regimens without PD-1/PD-L1 inhibitors.

Studies eligible for inclusion met the following criteria:

1. Cancer therapy randomized controlled clinical trial.
2. Experimental group: participants were treated with a single-agent PD-1/PD-L1 inhibitor, or PD-1/PD-L1 inhibitor plus chemotherapy.
3. Control group: participants were treated without PD-1/PD-L1 inhibitors.
4. Reported tabulated data on treatment-related toxicities.

Studies published online ahead of print were eligible, but meeting abstracts, reviews, commentaries, studies published only in abstract form, quality of life studies, cost effectiveness analyses, non-randomized trials, and those in which the toxicities of the drug could not be ascertained, such as when the control was a different dose of the same drug or another checkpoint inhibitor were excluded. Three investigators (K.X., C.L., and Z.L.) independently reviewed the list of retrieved articles to choose potentially relevant articles, and disagreements about studies were discussed and resolved with consensus. Three reviewers (K.X., C.L., and Z.L.) independently extracted data from studies and all discrepancies were resolved in consensus with all investigators. We extracted the authors, publication year, research institute, journal name, trial name, phase, cancer histotype, patient characteristics (sex and age, etc.), PD-1 and PD-L1 inhibitor used, dose escalation, dosing schedule, the sizes of intervention and control groups, number of all toxicities, criteria for toxicity reporting, median treatment time, and median follow-up from each study included. All-grade (severity) toxicity and grade 3 or higher (severity) toxicity data were both extracted.

### 2.3 Endpoint Setting and Stratification Strategy

Our primary outcome was the incidence of commonly described organ and system specific treatment-related toxicities. We used information from the publication and recorded data on toxicities. For some ambiguous data, we directly contacted study authors or pharmaceutical sponsors for additional information. We used the Common Terminology of Clinical Toxicities (CTCAE) categorization to identify Grades 3∼4 as severe/life threatening toxicity and CTCAE Grades 1∼2 as mild/moderate toxicity. If the study did not report a specific toxicity, we assumed that the event did not occur. Data from different dosing arms within the same study were extracted and reported separately.

The stratification strategy we adopted for the subgroup analysis include the following:

1. Subgroup analysis by toxicity grade (severity). As mentioned, the grade could be generally divided into two categories, the mild/moderate toxicity (Grade 1∼2) and severe/life-threatening toxicity (Grade 3∼4). We analyzed all the toxicities as a whole, as well as the Grade 3∼4 toxicities.
2. Subgroup analysis by specific system and organ. All toxicities together, constitutional toxicity, respiratory toxicity, cutaneous toxicity, gastrointestinal toxicity, hepatotoxicity, hematologic toxicity, neurologic toxicity, endocrine toxicity, inflammatory conditions of cardiac and skeletal muscle (myocarditis and myositis), urinary toxicity, and treatment event leading to discontinuation were analyzed, respectively.
3. Subgroup analysis by treatment regimens in the intervention arm. It should be noted that according to the treatment regimen of the intervention arm, two kinds of randomized control clinical trials were included. One is single agent PD-1/PD-L1 inhibitors, and one is PD-1/PD-L1 inhibitors plus chemotherapy. These two cases in comparison with the control were subgroup-analyzed.
4. Subgroup analysis by treatment regimens in the control arm, which could also be divided into two categories: the first is chemotherapy, and the second is CTLA-4 inhibitor, which would be analyzed separately also.
5. Subgroup analysis by drug type. Among all the included studies using PD-1 inhibitors, nivolumab and pembrolizumab were analyzed by subgroup. Because the numbers of included studies that using atezolizumab, durvalumab, or avelumab were small, we analyzed all the PD-L1 inhibitors together, rather than separating them.
6. Subgroup analysis by cancer histotype. Lung cancer and melanoma, with enough included studies, were subgroup-analyzed. Only subgroups including more than two studies were considered.
7. We would use the median follow-up time in the intervention arm to indirectly reflect the PD-1/PD-L1 inhibitors exposure time and administered dose to make the corresponding subgroup analysis if the median follow-up time (month) was detected as a factor for the between-study variance in the meta-regression analysis.

### 2.4 Data Synthesis and Analysis

We calculated overall event rates by dividing the total number of patients across trials with a given toxicity by the total number at risk. We examined the number of events for each immune-related toxicity of interest to determine whether meta-analysis was feasible. For each included study, we calculated relative risks (RRs) and 95% confidence intervals (95% CIs) for event rates in the intervention arm compared with control based on the reported number of events and sample size. We used the I^2^ index and the Cochran Q statistic to examine heterogeneity across trials for each outcome [58]. The statistical heterogeneity among studies was evaluated using Cochran’s Q test and I^2^ statistic, with values of 25%, 50%, and 75% representing low, moderate, and high heterogeneity, respectively. If significant heterogeneity was not present (P>0.05), summary RRs were estimated with a fixed effects model using the inverse variance method. A random effects model using the inverse variance method was used to calculate summary RRs and 95% confidence interval if significant heterogeneity was present (P≤0.05) [59]. If substantial heterogeneity was detected, we performed multivariate meta-regression analyses to explore the proportion of between-study variance explained by year of publication, country, intervention-arm patient percentage, male-female ratio, Eastern Cooperative Oncology Group (ECOG) score, median age, race (White race percentage), median follow-up (month), median progression free survival (PFS, month), and median overall survival (OS, month). Studies were weighted based on the inverse of the variance of the effect estimate [60, 61]. An estimation of publication bias was evaluated by the Beggs funnel plot, in which the SE of log (OR) of each study was plotted against its log (OR). An asymmetrical plot suggests possible publication bias. Egger’s linear regression test assessed funnel plot asymmetry, a statistical approach to identify funnel plot asymmetry on the natural logarithm scale of the ORs. All reported P-values are 2-sided. A P value less than 0.05 was considered to indicate statistical significance. All analyses were performed with the Stata version 14.2 (StataCorp, College Station, TX, USA) [62, 63].

### 2.5 Risk of Bias Assessment and Quality Assessment

Study methodological quality was assessed by using the five-point Jadad ranking system, that evaluates quality of randomization, double-blinding, and the flow of patients (withdrawals and dropouts), a practice in agreement with other meta-analyses done in this context. A clinical trial could receive a Jadad score of between zero (poor methodological quality) and five (optimal methodological quality). Two authors (K.X., C.L.) independently assessed the quality of all articles included in the review using the Cochrane Risk of Bias Tool and used a weighted Cohen’s kappa coefficient (κ) to measure agreement [64]. To optimize relevance to immune-related toxicities we evaluated the risk of bias with regard to toxicity outcomes, not the efficacy outcomes the individual studies were primarily designed to assess. Differences were resolved by consensus with the third author (L.Z.).

### 2.6 Patient Involvement

No patients were involved in setting the research question or the outcome measures, nor were they involved in developing plans for design or implementation of the study. No patients were asked to advise on interpretation or writing up of results. There are no plans to disseminate the results of the research to study participants or the relevant patient community. It was not evaluated whether the studies included in the review had any patient involvement.

## 3 Results

### 3.1 Literature Search, Eligible Studies and Characteristics

Literature search and review of reference lists identified 460 relevant publications (**Figure 1** showed the study selection flowchart.). After screening and eligibility assessment, we included in the meta-analysis a total of 29 clinical trials involving 8067 patients, with dates of publication ranging from 2014 to 2019 (**Table 1**) [65-93]. Data from all eligible studies were obtained from published manuscripts. The involved cancer histotypes included melanoma (n = 10), lung cancer (n = 15), gastrointestinal cancer (n = 1), genitourinary cancer (n = 2), Recurrent or Metastatic head and neck squamous cell carcinoma (HNSCC) (n = 1). The PD-1 and PD-L1 inhibitors used included nivolumab (n = 11), pembrolizumab (n = 11), atezolizumab (n = 4), avelumab (n = 1), and durvalumab (n = 2). Among all the included studies, 22 studies’ intervention arms adopted a treatment regimen of only PD-1/PD-L1 inhibitors, and 7 studies adopted PD-1/PD-L1 inhibitors plus chemotherapy. Across the meta-analysis, 23 studies used regular chemotherapy as the control arm, while 6 studies used CTLA-4 inhibitors. All the toxicities or toxicities were divided into nine categories according to the affected system or organ: 29 studies reported the constituional toxicity [fatigue (29 studies), asthenia (24 studies), pyrexia (17 studies), headache (9 studies), peripheral edema (10 studies), electrolyte imbalance (1 studies)], 25 studies reported the respiratory toxicity [cough (9 studies), dyspnea (14 studies), pneumonitis (25 studies)], 26 studies reported the cutaneous toxicity [pruritus (23 studies), rash (26 studies), alopecia (14 studies), vitiligo (9 studies), dermatitis (7 studies)], 29 studies reported the gastrointestinal toxicity [nausea (29 studies), vomiting (24 studies), decreased appetite (27 studies), diarrhea (29 studies), constipation (19 studies), colitis (21 studies)], 16 studies reported the hepatotoxicity [increase in alanine aminotransferase (ALT) (16 studies), increase in aspartate aminotransferase (AST) (15 studies), increase in bilirubin (BIU) (7 studies), hepatitis (16 studies)], 25 studies reported the hematologic toxicity [leukopenia (12 studies), neutropenia (19 studies), thrombocytopenia (10 studies), anemia (25 studies), thrombosis (8 studies)], 19 studies reported the neurotoxic toxicity [myasthenia gravis (2 studies), arthralgia (19 studies), myalgia (13 studies)], 27 studies reported the endocrinal toxicity [hypothyroidism (27 studies), hyperthyroidism (22 studies), adrenal insufficiency (13 studies), hypophyisitis (15 studies)], 6 studies reported the inflammatory conditions of cardiac and skeletal muscle (myocarditis and myositis), 13 studies reported the urinary toxicity (nephritis), and lastly, 28 studies reported the treatment event leading to discontinuation.

**Table 1.**
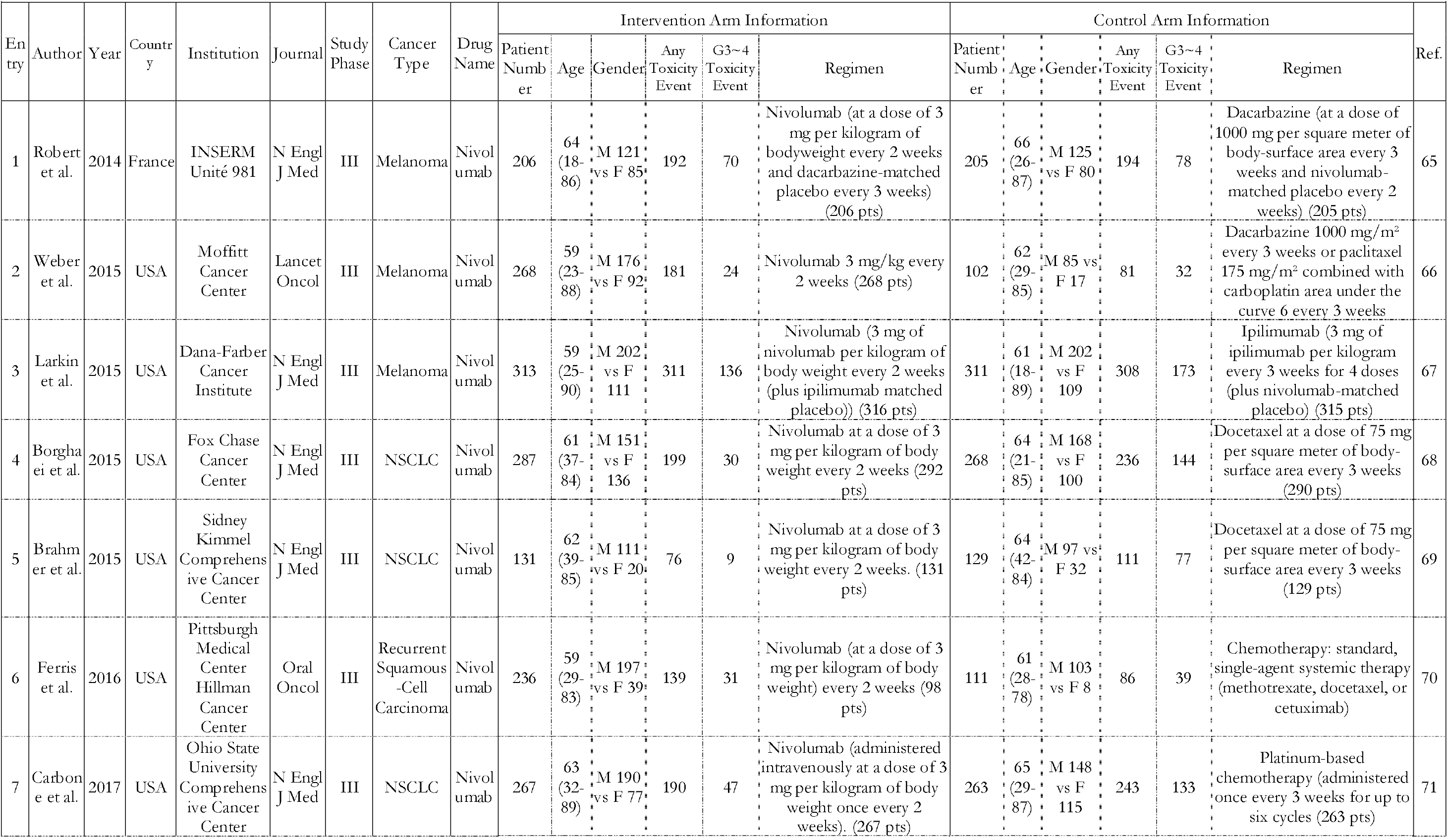
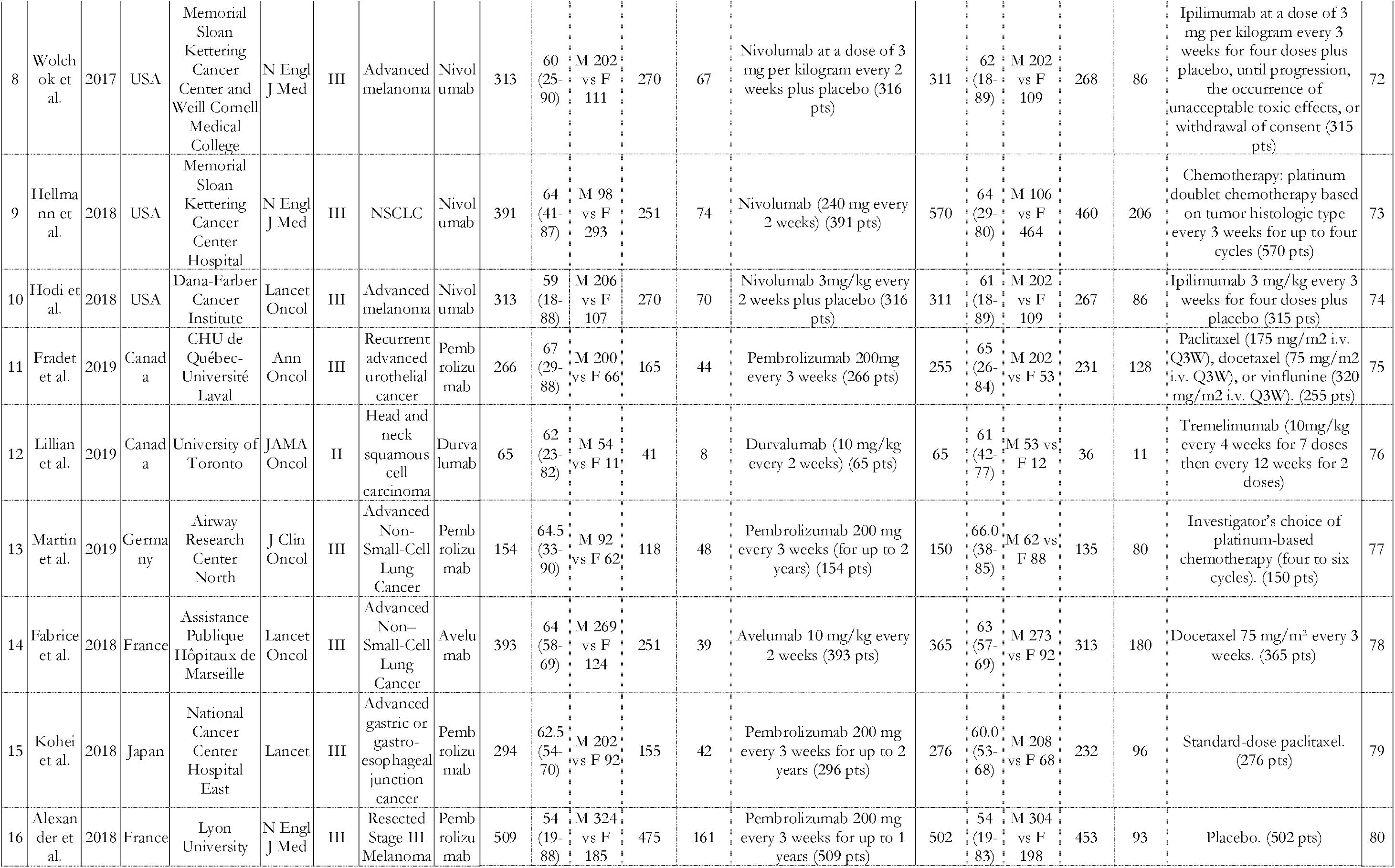
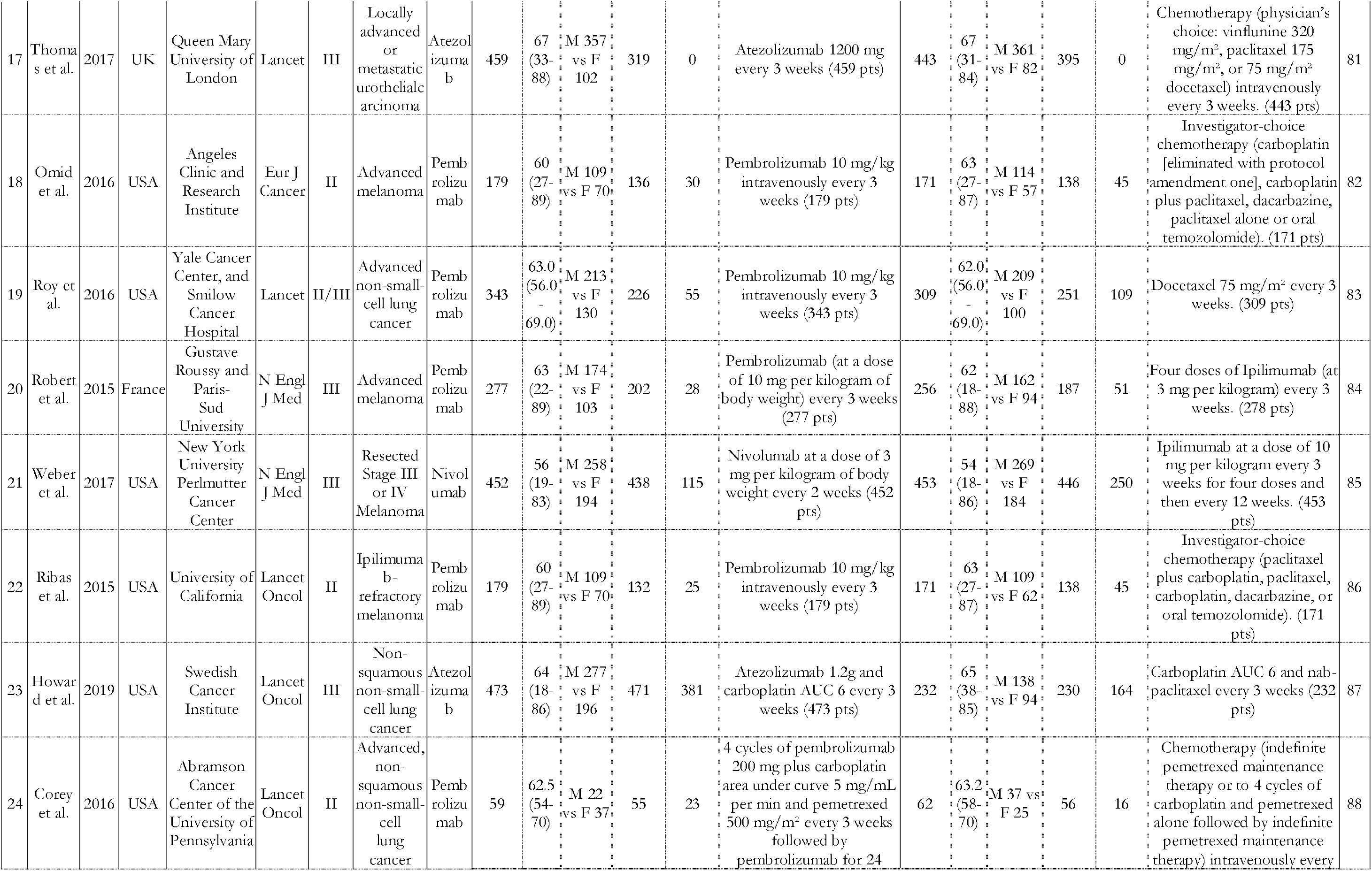
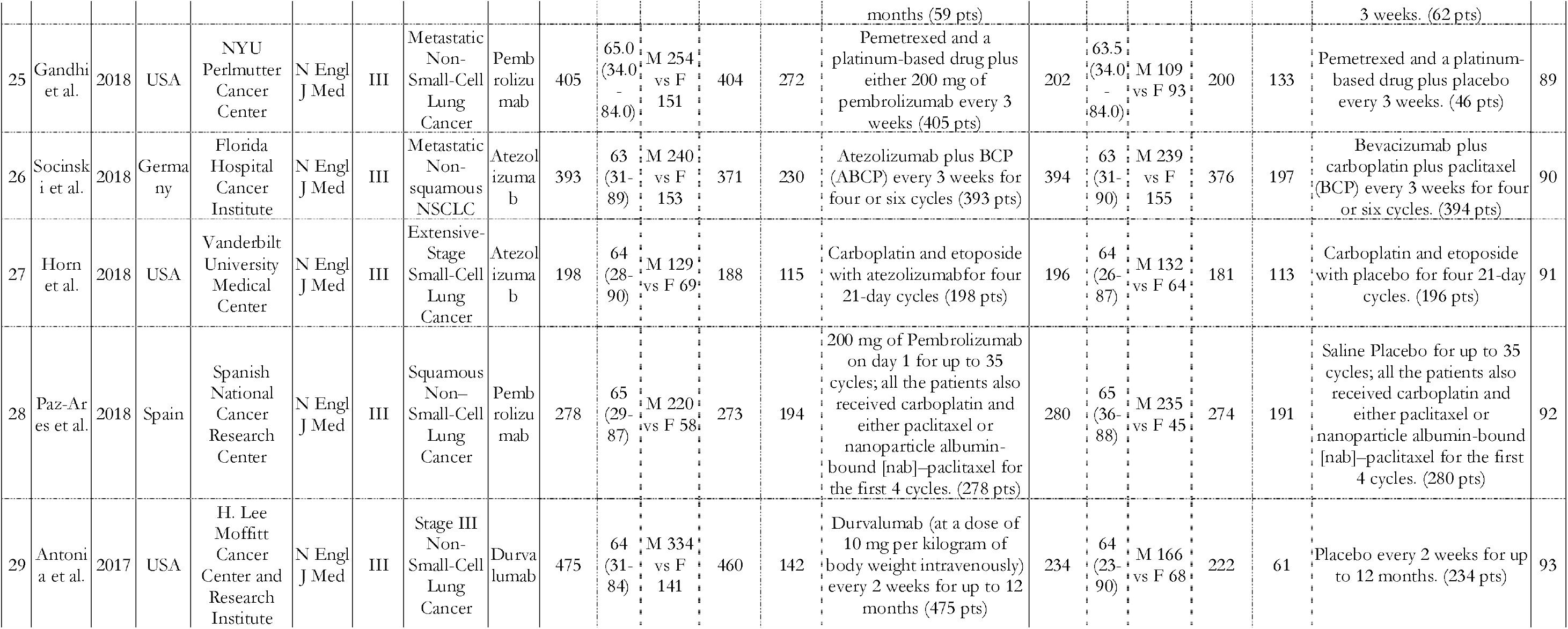
Characteristics of the Studies Included in This Meta-Analysis

**Figure 1.**
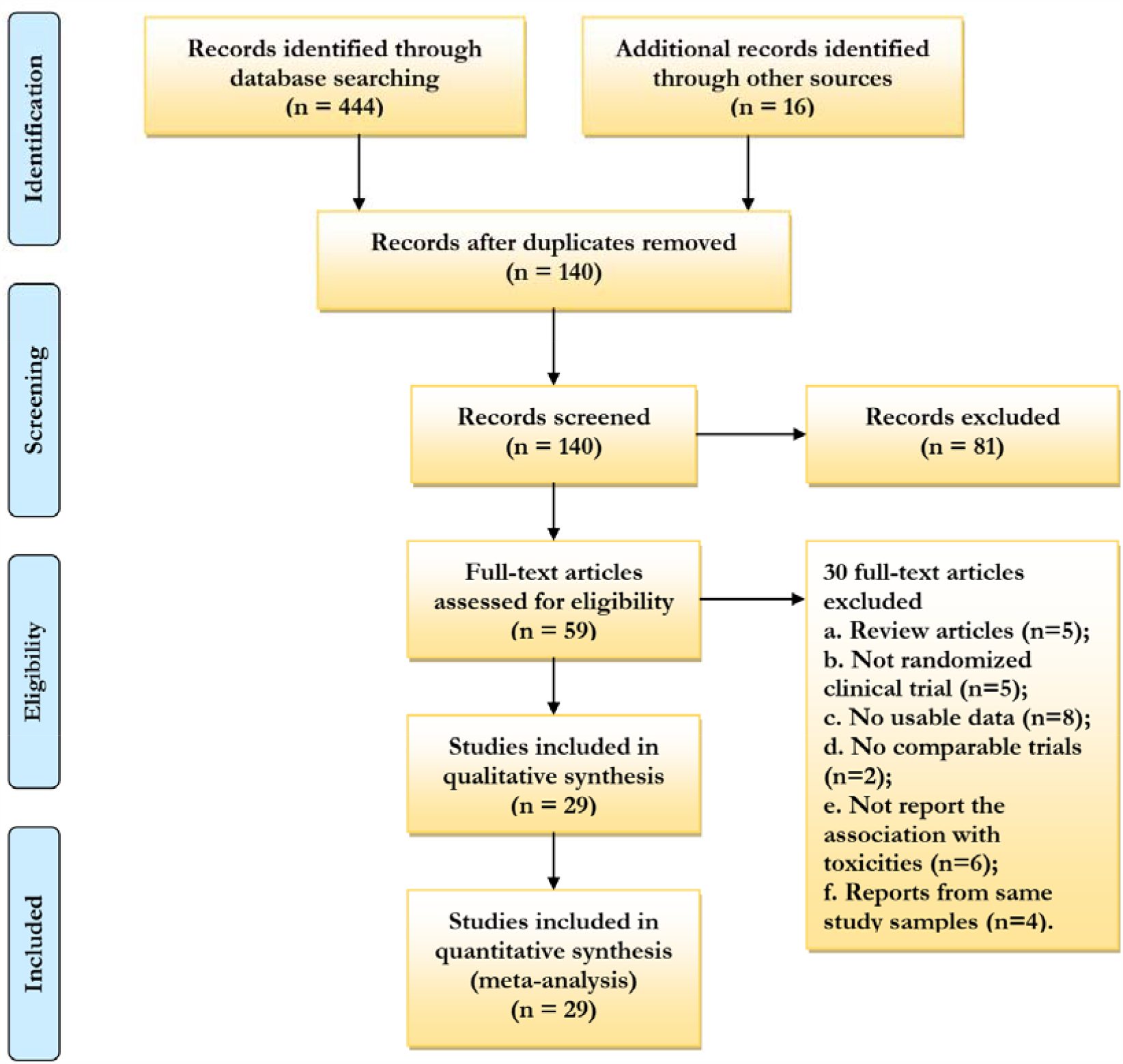
PRISMA Flow Diagram of Study Selection for this Meta-Analysis.

The numbers of patients in the intervention arm and control arm enrolled in each trial ranged between 59 and 509, 62 and 570, respectively. Of the total 16,173 patients included, 10,311 (63.75 %) were male and 5862 (36.25 %) were female. Of the total 8,576 patients included in the intervention arm, 5491 (64.03%) were male and 3085 (35.97%) were female. Of the total 7,597 patients included in the intervention arm, 4820 (63.45%) were male and 2777 (36.55%) were female. The median age of patients ranged from 54 to 67 years old across all included studies. 24 eligible studies were phase III studies, 5 studies were of phase II. 18 studies were conducted in the US, which accounted for the largest part, followed by 4 studies conducted in France, 2 studies conducted in Canada, 2 studies conducted in Germany, 1 study conducted in Japan, 1 study conducted in Spain, 1 study conducted in UK. There were 15 studies published in N Engl J Med, which accounted for the most, 6 in Lancet Oncol, and 1 in JAMA Oncol. Other involved journals included Lancet (3 study), Oral Oncol (1 studies), Ann Oncol (1 studies), Eur J Cancer (1 studies), and J Clin Oncol (1 studies). Toxicity was the primary endpoint for all the eligible clinical trials.

### 3.2 Incidence Proportion of Toxicities

The incidence proportion was defined as the number of patients with certain toxicities divided by the total number of patients in the corresponding intervention arm or control arm.

From **Table S2**, looking at any grade human system specific toxicities, among the 8576 total patients exposed to PD-1/PD-L1 inhibitors, 3626 (42.28%) had constitutional toxicities, 3626 (42.28%) had respiratory toxicities, 2627 (30.63%) had cutaneous toxicities, 4894 (57.07%) had gastrointestinal toxicities, 577 (6.73%) had hepatotoxicity, 2223 (25.92%) had hematologic toxicities, 783 (9.13%) had neurotoxic toxicities, 1017 (11.86%) had endocrine toxicities, 10 (0.12%) had inflammatory conditions of cardiac and skeletal muscle (myocarditis and myositis), 30 (0.35%) had urinary toxicities (nephritis), and 991 (11.56%) encountered treatment events leading to discontinuation. The most common Grade 3∼4 toxicities were hematologic toxicity which occurred in 807 (9.41%) patients, followed by gastrointestinal toxicity in 444 (5.18%) patients, and then constitutional toxicity in 226 (2.64%) patients. Among the 7597 total patients treated in control arms, 3506 (46.15%) had constitutional toxicities, 708 (9.32%) had respiratory toxicities, 2479 (32.63%) had cutaneous toxicities, 6043 (79.54%) had gastrointestinal toxicities, 465 (6.12%) had hepatotoxicity, 3165 (41.66%) had hematologic toxicities, 705 (9.28%) had neurotoxic toxicities, 324 (4.26%) had endocrine toxicities, 2 (0.03%) had inflammatory conditions of cardiac and skeletal muscle (myocarditis and myositis), 24 (0.32%) had urinary toxicities (nephritis), and 1124 (14.80%) encountered treatment events leading to discontinuation. The most common Grade 3∼4 toxicity for patients treated in control arms was hematologic toxicity which occurred in 1358 (17.88%) patients. **Figure 2** showed intuitively the incidence proportions of all toxicities at any grade and Grade 3∼4 based on the affected systems or organs for both the intervention arm and the control arm.

**Figure 2.**
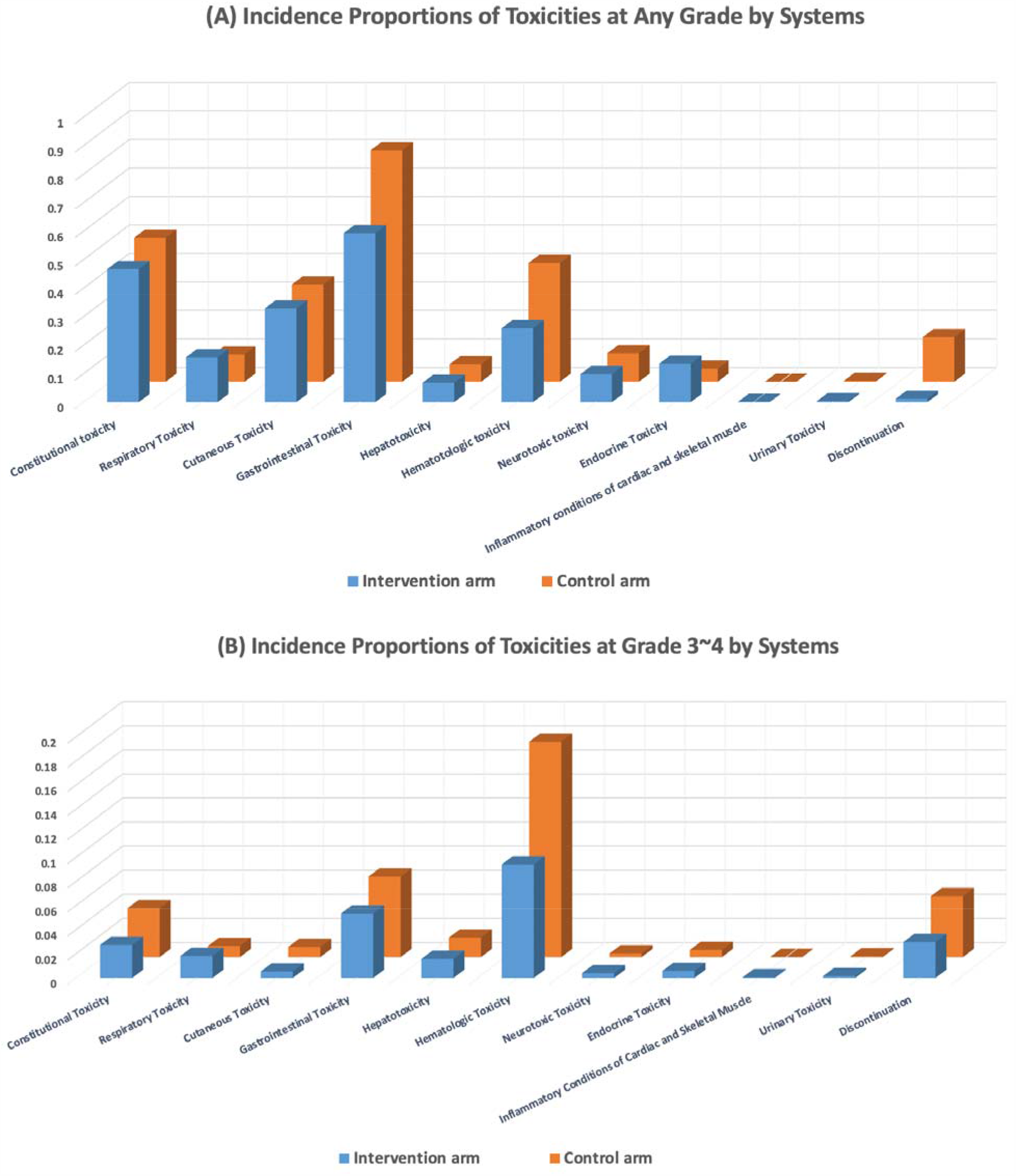
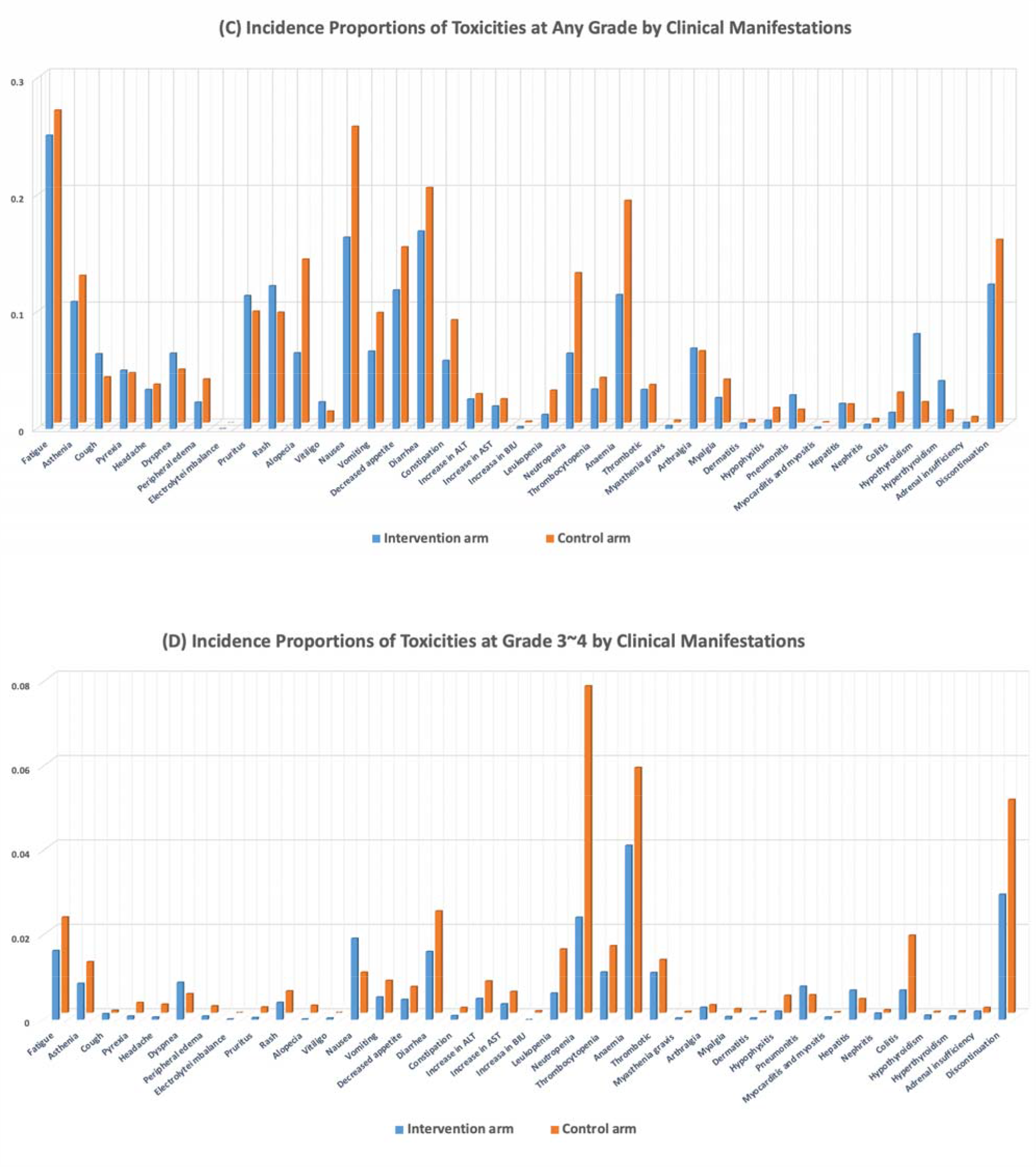
Incidence Proportions of All Toxicities at Any Grade and Grade 3∼4 Based on The Affected Systems or Organs for Both the Intervention Arm and the Control Arm. (**A**) Incidence Proportions of Toxicities at Any Grade by Systems; (**B**) Incidence Proportions of Toxicities at Grade 3∼4 by Systems; C) Incidence Proportions of Toxicities at Any Grade by Clinical Manifestations; (**D**) Incidence Proportions of Toxicities at Grade 3∼4 by Clinical Manifestations.

### 3.3 Overall Comparison of Toxicities

For all toxicities of any type, patients treated with PD-1/PD-L1 inhibitors were less likely to experience toxicities (all grades: RR 0.91, 95% CI 0.89-0.92; Grade 3∼4: RR 0.76, 95% CI 0.74-0.78) (**Figure 3**). For toxicities at any grade, at the human system level, when compared with patients treated in control arms, those treated with PD-1/PD-L1 inhibitors were at lower risks for gastrointestinal toxicity (RR 0.68, 95% CI 0.60-0.77), hematologic toxicity (RR 0.66, 95% CI 0.51-0.85), and treatment event leading to discontinuation (RR 0.78, 95% CI 0.72-0.84); and were at higher risks for respiratory toxicity (RR 1.74, 95% CI 1.33-2.28). At the organ level, those treated with PD-1/PD-L1 inhibitors were at lower risks for fatigue (RR 0.93), asthenia (RR 0.83), peripheral edema (RR 0.61), alopecia (RR 0.46), nausea (RR 0.63), vomiting (RR 0.70), decreased appetite (RR 0.78), diarrhea (RR 0.83), constipation (RR 0.66), leukopenia (RR 0.44), neutropenia (RR 0.50), anemia (RR 0.60), myalgia (RR 0.72), hypophyisitis (RR 0.46), colitis (RR 0.46); and were at higher risks for pyrexia (RR 1.17), cough (RR 1.63), dyspnea (RR 1.40), pruritus (RR 1.16), rash (RR 1.29), vitiligo (RR 2.34), increase in ALT (RR 1.03), increase in BIU (RR 1.88), thrombotic (RR 1.03), myasthenia Gravis (RR 1.51), arthralgia (RR 1.14), dermatitis (RR 2.00), pneumonitis (RR 2.48), myocarditis and myositis (RR 4.40), hepatitis (RR 1.31), nephritis (RR 1.10), hypothyroidism (RR 4.57), hyperthyroidism (RR 3.53), adrenal insufficiency (RR 1.09).

**Figure 3.**
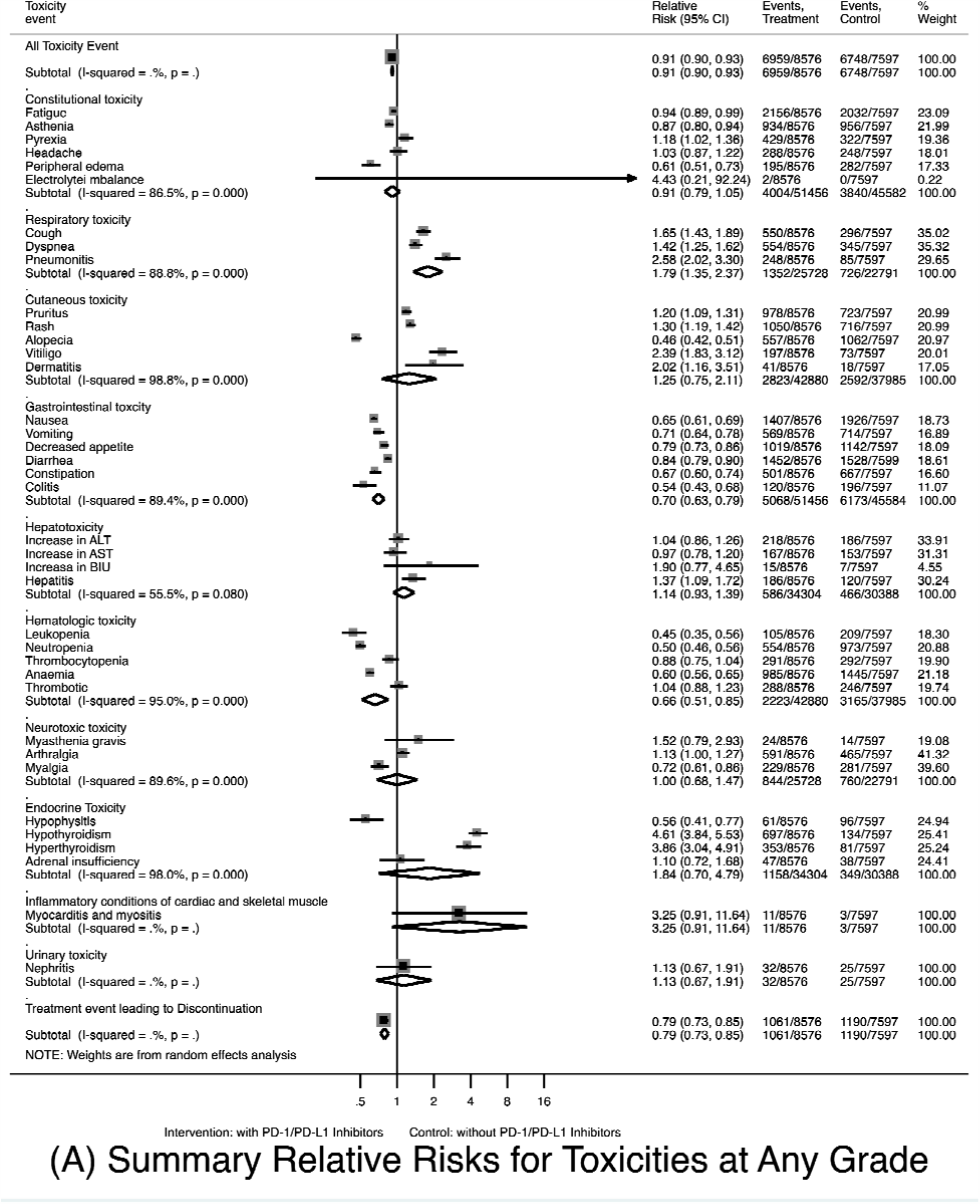
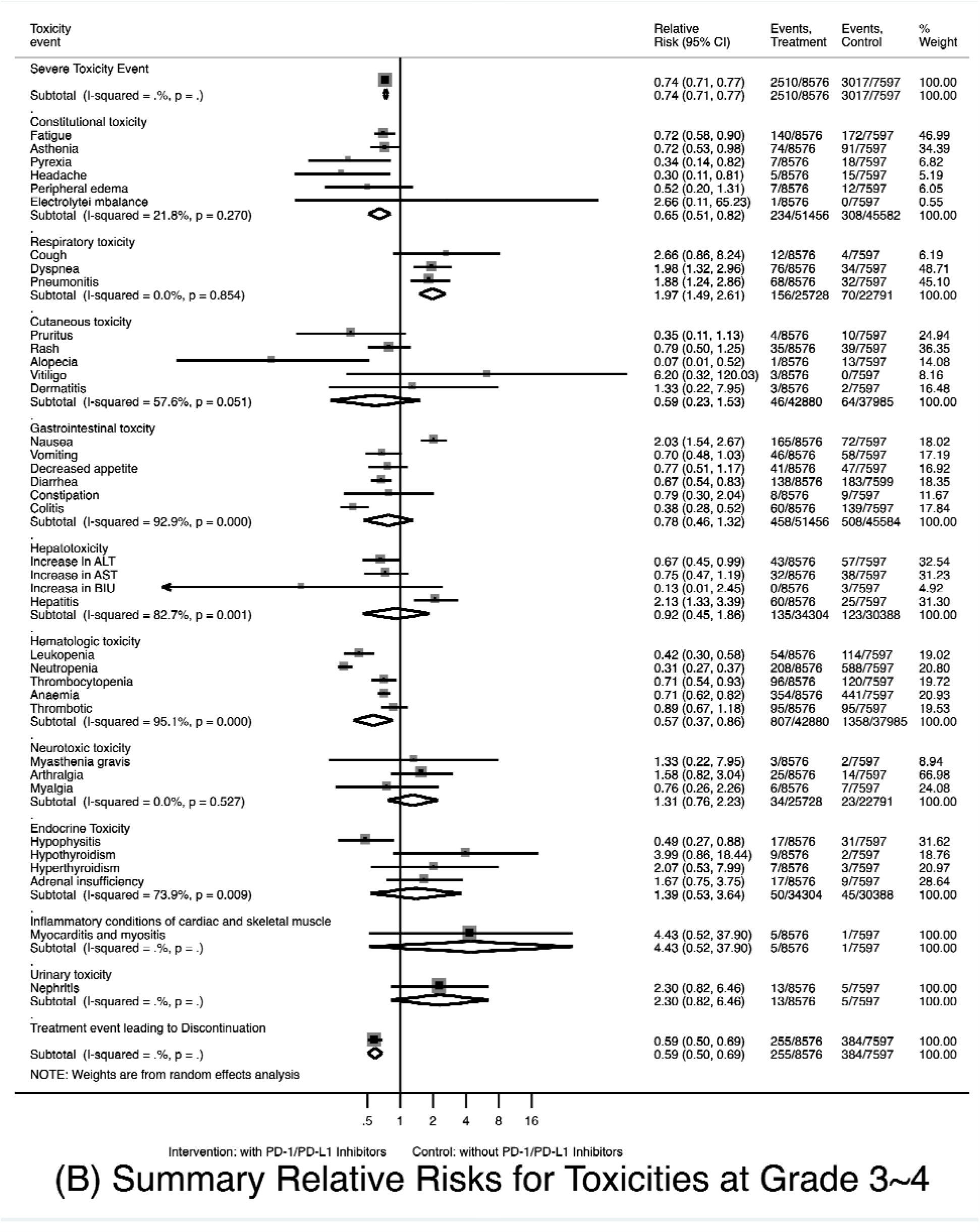
Forest Plots for the Overall Comparison of Toxicities. (**A**) Summary Relative Risks for Toxicities at Any Grade; (**B**) Summary Relative Risks for Toxicities at Grade 3∼4.

When evaluating Grade 3∼4 toxicities specifically and at the organ system level, compared with patients treated in control arms, those treated with PD-1/PD-L1 inhibitors were at lower risks for constitutional toxicity (RR 0.63, 95% CI 0.51-0.78), hepatotoxicity (RR 0.55, 95% CI 0.37-0.83), hematologic toxicity (RR 0.55, 95% CI 0.37-0.83), and treatment event leading to discontinuation (RR 0.58, 95% CI 0.49-0.67); and were at higher risks for respiratory toxicity (RR 1.92, 95% CI 1.45-2.55). Those treated with PD-1/PD-L1 inhibitors were at lower risks for fatigue (RR 0.69), asthenia (RR 0.68), pyrexia (RR 0.35), headache (RR 0.29), peripheral Edema (RR 0.51), pruritus (RR 0.36), rash (RR 0.79), alopecia (RR 0.06), vomiting (RR 0.68), decreased Appetite (RR 0.74), diarrhea (RR 0.63), constipation (RR 0.76), increase in ALT (RR 0.67), increase in AST (RR 0.74), increase in BIU (RR 0.13), leukopenia (RR 0.41), neutropenia (RR 0.31), thrombocytopenia (RR 0.70), anemia (RR 0.66), thrombotic (RR 0.88), myalgia (RR 0.75), hypophyisitis (RR 0.40), colitis (RR 0.32); and at higher risks for cough (RR 2.71), dyspnea (RR 1.97), nausea (RR 1.76), myasthenia gravis (RR 10.54), arthralgia (RR 1.39), dermatitis (RR 1.32), pneumonitis (RR 1.78), myocarditis and myositis (RR 3.52), hepatitis (RR 1.95), nephritis (RR 1.93), hypothyroidism (RR 4.21), hyperthyroidism (RR 1.81), adrenal insufficiency (RR 1.56).

### 3.4 Quality and Heterogeneity of Included Studies

Randomized treatment allocation sequences were generated in all studies. The method quality of the included trials was all good. The risks of bias of 29 included randomized controlled trials were shown in **Table 2**. The only issue affecting quality was lack of blinding because all trials were open labelled. From the forest plot of the toxicity at Any Grade, constitutional toxicity, respiratory toxicity, cutaneous toxicity, gastrointestinal toxicity, hematologic toxicity, neurotoxic toxicity, endocrinal toxicity had high heterogeneity (all I^2^>75%, P<0.001) and hepatotoxicity with moderate heterogeneity (I^2^=55.5%, P=0.08). In toxicities at Grade 3∼4, gastrointestinal toxicity, hepatotoxicity, hematologic toxicity, endocrinal toxicity had high heterogeneity (all I^2^>75%, P<0.05), cutaneous toxicity with moderate heterogeneity (I^2^=59.3%, P=0.044), constitutional toxicity (I^2^=21.8%, P=0.27), respiratory toxicity (I^2^=0%, P=0.854) and neurotoxic toxicity(I^2^=0%, P=0.527) with low heterogeneity.

**Table 2.** **Risk of Bias of Included Randomized Controlled Trials**

| Author | Year | Randomization | Allocation Concealment | Blinding of Participants and Staff | Blinding of Outcome Assessors* | Incomplete Outcome Data† | Selective Outcome Reporting† | Other Sources of Bias |
| --- | --- | --- | --- | --- | --- | --- | --- | --- |
| Robert et al. | 2014 | Low | Low | Low | Low | Low | Low | Low |
| Weber et al. | 2015 | Low | Low | Low | Low | Low | Low | Low |
| Larkin et al. | 2015 | Low | Low | Low | Low | Low | Low | Low |
| Borghaei et al. | 2015 | Low | Low | Low | Low | Low | Low | Low |
| Brahmer et al. | 2015 | Low | Low | Low | Low | Low | Low | Low |
| Robert et al. | 2015 | Low | Low | Low | Low | Low | Low | Low |
| Ribas et al. | 2015 | Low | Low | Low | Low | Low | Low | Low |
| Ferris et al. | 2016 | Low | Low | Low | Low | Low | Low | Low |
| Omid et al. | 2016 | Low | Low | Low | Low | Low | High | Low |
| Roy et al. | 2016 | Low | Low | Low | Low | Low | Low | Low |
| Corey et al. | 2016 | Low | Low | Low | High | Low | Low | Low |
| Carbone et al. | 2017 | Low | Low | Low | Low | Low | Low | Low |
| Wolchok et al. | 2017 | Low | Low | Low | Low | Low | Low | Low |
| Thomas et al. | 2017 | Low | Low | Low | Low | Low | Low | Low |
| Weber et al. | 2017 | Low | Low | Low | Low | Low | Low | Low |
| Antonia et al. | 2017 | Low | Low | Low | Low | Low | Low | Low |
| Hellmann et al. | 2018 | Low | Low | Low | Low | Low | Low | Low |
| Hodi et al. | 2018 | Low | Low | Low | Low | Low | Low | Low |
| Fabrice et al. | 2018 | Low | Low | Low | Low | Low | Low | Low |
| Kohei et al. | 2018 | Low | Low | Low | Low | High | Low | Low |
| Alexander et al. | 2018 | Low | Low | Low | Low | Low | Low | Low |
| Gandhi et al. | 2018 | Low | Low | Low | Low | Low | Low | Low |
| Socinski et al. | 2018 | Low | Low | Low | Low | Low | Low | Low |
| Horn et al. | 2018 | Low | Low | Low | Low | Low | Low | Low |
| Paz-Ares et al. | 2018 | Low | Low | Low | Low | Low | Low | Low |
| Fradet et al. | 2019 | Low | Low | Low | Low | Low | Low | Low |
| Lillian et al. | 2019 | Low | Low | Low | Low | High | Low | Low |
| Martin et al. | 2019 | Low | Low | Low | Low | Low | Low | Low |
| Howard et al. | 2019 | Low | Low | Low | Low | Low | Low | Low |

1) \*Based on clinician blinding since clinicians assessed toxicities for all studies.
2) †Applies to adverse events.

### 3.5 Subgroup Analysis by Treatment Regimens in the Intervention Arm

There were two kinds of treatment regimens for the intervention arms: 1) single agent PD-1/PD-L1 inhibitors (22 studies), and 2) PD-1/PD-L1 inhibitors plus chemotherapy (7 studies, all were for lung cancer treatment and the regimens in corresponding control arms were all chemotherapy). In order to avoid the interference effects of the chemotherapy in the intervention arm on the overall results, a subgroup analysis was performed. Patients treated with single PD-1/PD-L1 inhibitors were less likely to experience overall toxicities of any type than their counterparts in the control arms (all grades: RR 0.86, 95% CI 0.80-0.92; Grade 3∼4: RR 0.56, 95% CI 0.42-0.75). System- and organ-specifically, they were less likely to experience constitutional toxicity (all grades: RR 0.75, 95% CI 0.66-0.84; Grade 3∼4: RR 0.39, 95% CI 0.30-0.51), including fatigue, asthenia, and headache at both all grades and Grade 3∼4and pyrexia only at Grade 3∼4 (RR 0.26, 95% CI 0.09-0.75), peripheral Edema only at any grade (all grades: RR 0.14, 95% CI 0.05-0.41); more likely for overall respiratory toxicity (all grades: RR 1.61, 95% CI 1.24-2.09; Grade 3∼4: RR 1.99, 95% CI 1.28-3.09), especially for pneumonitis (all grades: RR 2.40, 95% CI 1.32-4.38; Grade 3∼4: RR 2.16, 95% CI 1.12-4.15); more likely for cutaneous toxicity at any grade (RR 1.28, 95% CI 1.00-1.64), but less likely at Grade 3∼4 (RR 0.55, 95% CI 0.35-0.86); less likely for gastrointestinal toxicity (all grades: RR 0.51, 95% CI 0.45-0.57; Grade 3∼4: RR 0.36; 95% CI 0.27-0.47), including nausea, vomiting, decreased appetite, diarrhea, and constipation for both all grades and Grade 3∼4; less likely for hematologic toxicity (all grades: RR 0.11, 95% CI 0.08-0.16; Grade 3∼4: RR 0.07, 95% CI 0.05-0.11), including leukopenia, neutropenia, thrombocytopenia, anemia, and thrombotic at both all grades and Grade 3∼4; more likely for endocrine toxicity at any grade (RR 2.99, 95% CI 2.10-4.26), including hypothyroidism, hyperthyroidism, adrenal insufficiency and hypophyisitis at any grade; and less likely for treatment event leading to discontinuation (all grades: RR 0.60; 95% CI 0.46-0.77; Grade 3∼4: RR 0.68; 95% CI 0.38-1.20). There was no significant difference for hepatotoxicity, neurotoxic toxicity, myocarditis and myositis, and nephritis (all P>0.05).

Compared with patients treated in control arms, those treated with PD-1/PD-L1 inhibitors (plus chemotherapy) were at higher risk for overall toxicities of any type (all grades: RR 1.01, 95% CI 1.00-1.01; Grade 3∼4: RR 1.01, 95% CI 1.00-1.02), including constitutional toxicity (all grades: RR 1.11, 95% CI 1.01-1.21; Grade 3∼4: RR 1.13, 95% CI 0.87-1.48), cutaneous toxicity (all grades: RR 1.47, 95% CI 1.25-1.72; Grade 3∼4: RR 1.16, 95% CI 0.51-2.64), gastrointestinal toxicity (all grades: RR 1.15, 95% CI 1.06-1.26; Grade 3∼4: RR 1.61, 95% CI 1.15-2.27), hepatotoxicity (all grades: RR 1.40, 95% CI 1.10-1.77), hematologic toxicity (all grades: RR 1.14, 95% CI 1.03-1.26; Grade 3∼4: RR 1.10, 95% CI 1.00-1.22), neurotoxic toxicity (all grades: RR 1.20, 95% CI 1.04-1.39), endocrine toxicity (all grades: RR 3.22, 95% CI 2.22-4.69), and treatment event leading to discontinuation (all grades: RR 1.42, 95% CI 1.16-1.74; Grade 3∼4: RR 1.65, 95% CI 1.10-2.47).

### 3.6 Subgroup Analysis by Treatment Regimens in the Control Arm

There were also two kinds of treatment regimen for the control arms: 1) chemotherapy (23 studies), and 2) CTLA-4 inhibitor targeted therapy (6 studies). Considering the possible toxicity differences between conventional chemotherapies and CTLA-4 inhibitors, sub-group analysis was conducted herein to decrease the cross confounding and chiasma interference. Compared with patients treated by conventional chemotherapies in control arms, patients treated with PD-1/PD-L1 inhibitors were at lower risks to have overall toxicities of any type (all grades: RR 0.87, 95% CI 0.81-0.93; Grade 3∼4: RR 0.63, 95% CI 0.52-0.75). Specifically, they were at lower risks for constitutional toxicity (all grades: RR 0.81, 95% CI 0.73-0.90; Grade 3∼4: RR 0.57, 95% CI 0.43-0.75), including fatigue at both all grades and Grade 3∼4, and asthenia and peripheral edema at any grade; at higher risks for pneumonitis (all grades: RR 2.85, 95% CI 1.89-4.31); at higher risks for cutaneous toxicity (all grades: RR 1.54, 95% CI 1.22-1.95), including pruritus, rash, and vitiligo at any grade; at lower risks for gastrointestinal toxicity at any grade (RR 0.65, 95% CI 0.58-0.74), including nausea, vomiting, decreased appetite, and constipation at any grade, but not for colitis (all grades: RR 2.69, 95% CI 1.29-5.61; Grade 3∼4: RR 2.16, 95% CI 1.16-4.02); at higher risks for hepatotoxicity of any type (all grades: RR 1.40, 95% CI 1.18-1.85; Grade 3∼4: RR 1.52, 95% CI 1.00-2.31), including increase in ALT, AST and BIU, hepatitis at any grade and grade 3∼4, but not for increase in BIU at Grade 3∼4 (RR 0.16, 95% CI 0.01-4.02); at lower risks for hematologic toxicity (all grades: RR 0.27, 95% CI 0.21-0.35; Grade 3∼4: RR 0.28, 95% CI 0.21-0.39), including leukopenia, thrombocytopenia, and anemia at both all grades and Grade 3∼4, and neutropenia at any grade; at lower risks for neurotoxic toxicity (all grades: RR 0.75, 95% CI 0.57-0.99), but not for neurotoxic toxicity at Grade 3∼4 (RR 1.02, 95% CI 0.55-1.88), mainly myalgia (all grades: RR 0.54, 95% CI 0.29-0.98); at higher risks for endocrine toxicity (all grades: RR 4.64, 95% CI 3.58-6.02; Grade 3∼4: RR 2.41, 95% CI 1.38-4.27), including hypothyroidism and hyperthyroidism at any grade, and hypophyisitis at both any grade (RR 4.34, 95% CI 2.20-8.59) and Grade 3∼4 (RR 3.26, 95% CI 1.07-9.89). No significant difference was found for respiratory toxicity, myocarditis and myositis, nephritis, and treatment event leading to discontinuation (all P>0.05).

Compared with patients treated by CTLA-4 inhibitors in control arms, patients in intervention arms were more likely to experience fatigue (all grades: RR 1.20, 95% CI 1.08-1.34), asthenia (all grades: RR 1.31, 95% CI 1.03-1.67), less likely to experience headache (all grades: RR 0.76, 95% CI 0.58-0.98; Grade 3∼4: RR 0.22, 95% CI 0.06-0.89), pruritus (all grades: RR 0.61, 95% CI 0.54-0.69), rash (Grade 3∼4: RR 0.28, 95% CI 0.14-0.58), nausea (all grades: RR 0.82, 95% CI 0.69-0.97), diarrhea (all grades: RR 0.59, 95% CI 0.53-0.66; Grade 3∼4: RR 0.31, 95% CI 0.19-0.49), colitis (all grades: RR 0.23, 95% CI 0.15-0.34; Grade 3∼4: RR 0.14, 95% CI 0.08-0.27), hypophyisitis (all grades: RR 0.17, 95% CI 0.10-0.29; Grade 3∼4: RR 0.19, 95% CI 0.08-0.44), and treatment event leading to discontinuation (all grades: RR 0.52, 95% CI 0.30-0.91; Grade 3∼4: RR 0.37, 95% CI 0.18-0.76). No significant difference was found for respiratory toxicity, hepatotoxicity, hematologic toxicity, inflammatory conditions of cardiac and skeletal muscle, and urinary toxicity (all P>0.05).

### 3.7 Subgroup Analysis by Drug Type in the Intervention Arm

Compared with patients in control arms, patients treated with nivolumab were less likely to experience constitutional toxicity (all grades: RR 0.88, 95% CI 0.80-0.95; Grade 3∼4: RR 0.57, 95% CI 0.39-0.81), including fatigue, asthenia, pyrexia, and headache at Grade 3∼4; more likely for overall respiratory toxicity (all grades: RR 1.27, 95% CI 1.02-1.58; Grade 3∼4: RR 1.94, 95% CI 1.16-3.24); more likely for overall cutaneous toxicity at any grade (all grades: RR 1.31, 95% CI 1.00-1.72) (pruritus: all grades RR 1.73, 95% CI 1.07-2.81; vitiligo: all grades RR 2.37, 95% CI 1.35-4.18), but less likely for Grade 3∼4 (RR 0.53, 95% CI 0.32-0.88); less likely for gastrointestinal toxicity (all grades: RR 0.49, 95% CI 0.42-0.57; Grade 3∼4: RR 0.28, 95% CI 0.21-0.37), including nausea, vomiting, decreased appetite, diarrhea, and colitis at both any grade and Grade 3∼4, and constipation only at any grade (RR 0.38, 95% CI 0.19-0.78); less likely for hematologic toxicity (all grades: RR 0.12, 95% CI 0.07-0.20; Grade 3∼4: RR 0.07, 95% CI 0.04-0.13), including leukopenia, neutropenia, thrombocytopenia, and anemia at both all grades and Grade 3∼4; more likely for endocrine toxicity at any grade (all grades: RR 1.98, 95% CI 1.29-3.06), including hypothyroidism, hyperthyroidism at any grade (but less likely for hypophyisitis at all grades: RR 0.18, 95% CI 0.10-0.32); and less likely for treatment event leading to discontinuation (all grades: RR 0.52, 95% CI 0.36-0.76; Grade 3∼4: RR 0.53, 95% CI 0.29-0.95).

The overall comparison spectrum for pembrolizumab was similar to that for nivolumab, except for the following points: 1) patients treated with pembrolizumab were more likely to experience rash (all grades: RR 1.80, 95% CI 1.40-2.31); 2) they were more likely to develop hepatitis (all grades: RR 3.62, 95% CI 1.37-9.54; Grade 3∼4: RR 5.80, 95% CI 1.70-19.76); 3) they were more likely to develop pneumonitis (all grades: RR 3.22, 95% CI 2.08-4.96; Grade 3∼4: RR 2.32, 95% CI 1.20-4.48); 4) there was no significant difference for vitiligo, diarrhea, constipation, and colitis (all P>0.05).

The overall comparison spectrum for PD-L1 inhibitors was also similar to PD-1 inhibitors, except for: 1) patients treated with PD-L1 inhibitors were more likely to experience cough at any grade (All grades: RR 1.47, 95% CI 1.20-1.79; Grade 3∼4: RR 1.85, 95% CI 0.29-11.88); 2) They were more likely to experience colitis at any grade (all grades: RR 3.58, 95% CI 1.12-11.45; Grade 3∼4: RR 3.03, 95% CI 0.83-11.06); 3) there was no significant difference for gastrointestinal toxicity (all grades: RR 0.84, 95% CI 0.69-1.03; Grade 3∼4: RR 1.43, 95% CI 0.81-2.53), and leukopenia (all grades: RR 0.85, 95% CI 0.31-2.34; Grade 3∼4: RR 0.94, 95% CI 0.24-3.73).

### 3.8 Subgroup Analysis by Cancer Histotype

The toxicity spectrum was different between patients with melanoma and lung cancer. For melanoma, compared with patients in control arms, patients treated with PD-1/PD-L1 inhibitors were at lower risks to have headache (all grades: RR 0.73, 95% CI 0.59-0.91; Grade 3∼4: RR 0.22, 95% CI 0.06-0.89), rash at Grade 3∼4 (RR 0.37, 95% CI 0.19-0.73); nausea (all grades: RR 0.60, 95% CI 0.42-0.85; Grade 3∼4: RR 0.26, 95% CI 0.10-0.67), vomiting at any grade (all grades: RR 0.47, 95% CI 0.31-0.73), decreased appetite at any grade (all grades: RR 0.71, 95% CI 0.58-0.88), diarrhea (all grades: RR 0.77, 95% CI 0.62-0.95; Grade 3∼4: RR 0.42, 95% CI 0.26-0.69), colitis at Grade 3∼4 (RR 0.31, 95% CI 0.11-0.84); leukopenia (all grades: RR 0.09, 95% CI 0.03-0.25; Grade 3∼4: RR 0.09, 95% CI 0.02-0.37), neutropenia at Grade 3∼4 (RR 0.08, 95% CI 0.02-0.34), thrombocytopenia (all grades: RR 0.13, 95% CI 0.02-0.97; Grade 3∼4: RR 0.16, 95% CI 0.03-0.95), anemia at Grade 3∼4 (RR 0.10, 95% CI 0.03-0.33), hypophyisitis at Grade 3∼4 (RR 0.32, 95% CI 0.12-0.80); but were at higher risks to have asthenia at any grade (all grades: RR:1.14, 95% CI 1.00-1.29), dyspnea at any grade (all grades: RR 1.60, 95% CI 1.15-2.23), vitiligo at any grade (all grades: RR 2.69, 95% CI 1.54-4.68), dermatitis at any grade (all grades: RR 4.03, 95% CI 1.03-15.68), arthralgia at any grade (all grades: RR 1.28, 95% CI 1.00-1.63), hypothyroidism at any grade (all grades: RR 3.12, 95% CI 2.02-4.82), hyperthyroidism at any grade (all grades: RR 3.11, 95% CI 1.89-5.11), and hypophyisitis at any grade (all grades: RR 3.12, 95% CI 2.02-4.82).

For lung cancer, patients treated with PD-1/PD-L1 inhibitors were at lower risks to have fatigue at any grade (all grades: RR 0.80, 95% CI 0.67-0.96), asthenia at any grade (all grades: RR 0.71, 95% CI 0.58-0.87), nausea at any grade (all grades: RR 0.58, 95% CI 0.42-0.80), decreased appetite at any grade (all grades: RR 0.75, 95% CI 0.59-0.97), leukopenia (all grades: RR 0.30, 95% CI 0.12-0.74; Grade 3∼4: RR 0.07, 95% CI 0.01-0.55), neutropenia (all grades: RR 0.19, 95% CI 0.09-0.40; Grade 3∼4: RR 0.13, 95% CI 0.05-0.33), and anemia (all grades: RR 0.35, 95% CI 0.23-0.53; Grade 3∼4: RR 0.41, 95% CI 0.24-0.70); but were at higher risks to have pruritus at any grade (all grades: RR 3.33, 95% CI 2.20-5.03), rash at any grade (all grades: RR 1.93, 95% CI 1.60-2.34), increase in ALT at any grade (all grades: RR 1.38, 95% CI 1.04-1.81), increase in AST at any grade (all grades: RR 1.50, 95% CI 1.07-2.12), hypophysitis at any grade (all grades: RR 6.65, 95% CI 4.01-11.03), pneumonitis at any grade (all grades: RR 2.48, 95% CI 1.56-3.93; Grade 3∼4: RR 2.03, 95% CI 1.23-3.35), hepatitis at any grade (all grades: RR 1.53, 95% CI 1.03-2.29), colitis (all grades: RR 2.61, 95% CI 1.00-6.79; Grade 3∼4: RR 2.16, 95% CI 1.06-4.41), hypothyroidism at any grade (all grades: RR 6.65, 95% CI 4.01-11.03), and hyperthyroidism at any grade (all grades: RR 3.69, 95% CI 2.49-5.46).

### 3.9 Meta-Regression and Publication Bias Analysis

**Table 3** showed the results of the multivariate meta-regression for the included 29 studies. The between-study variance could not be explained by the year of publication, country, intervention-arm patient percentage, Eastern Cooperative Oncology Group (ECOG) score, median age, race (White percentage), median follow-up (<=18 months: 14 studies, RR=0.87, 95% CI 0.79∼0.96; >18 months: 15 studies, RR=0.92, 95% CI 0.87∼0.97; I^2^=91.3, τ2=0.013, and P=0.79), median progression free survival (PFS, month), or median overall survival (OS, month) (all P >0.05). Only male patient percentage was a statistically significant variable (I^2^=89.1, τ2=0.01, and P=0.001). Seeing this, the subgroup analysis based on the PD-1/PD-L1 inhibitors exposure time and administered dose reflected by the median follow-up time was waived. **Figure 4** showed that the funnel plot and Eggers statistical test indicated evidence of heterogeneities and/or publication bias in the studies included in the meta-analysis with scatters beyond 95% CI and asymmetry display (P<0.001).

**Table 3.** **Summary of Meta Regression for the Included 29 Clinical Trials with Any Toxicities**

| Regression Variables | Number of Studies | Random Effect Summary RR (95% CI) | I<sup>2</sup> (%) | τ<sup>2</sup> | P-value |
| --- | --- | --- | --- | --- | --- |
| <u>Year of Publication</u> |  |  |  |  |  |
| Before 2018 | 16 | 0.89 (0.84-0.96) | 90.3 | 0.14 | 0.78 |
| 2018 Onwards | 13 | 0.90 (0.86-0.94) |  |  |  |
| <u>Country</u> |  |  |  |  |  |
| USA | 18 | 0.92 (0.87-0.96) | 86.1 | 0.16 | 0.51 |
| Non-USA | 11 | 0.88 (0.79-0.97) |  |  |  |
| <u>Trial Patient Percentage</u> |  |  |  |  |  |
| 50% | 23 | 0.88 (0.84-0.93) | 90.1 | 0.01 | 0.29 |
| Non 50% | 6 | 0.95 (0.88-1.03) |  |  |  |
| <u>Male Percentage</u> |  |  |  |  |  |
| <=0.64<sup>^</sup> | 17 | 0.97 (0.95-1.00) | 89.1 | 0.01 | **0.001** |
| >0.64 | 12 | 0.80 (0.70-0.93) |  |  |  |
| <u>ECOG=0*</u> |  |  |  |  |  |
| <50% | 18 | 0.85 (0.78-0.92) | 90.4 | 0.011 | 0.06 |
| >=50% | 8 | 0.98 (0.94-1.02) |  |  |  |
| <u>Trial Median Age</u> |  |  |  |  |  |
| <=62.5 | 15 | 0.91 (0.86-0.97) | 90.5 | 0.01 | 0.66 |
| >62.5 | 14 | 0.89 (0.82-0.96) |  |  |  |
| <u>Race (White Percentage) *</u> |  |  |  |  |  |
| <=85% | 9 | 0.88 (0.79-0.98) | 91.3 | 0.01 | 0.79 |
| >85% | 5 | 0.90 (0.81-1.00) |  |  |  |
| <u>Median Follow-up (Month)</u> |  |  |  |  |  |
| <=18 | 14 | 0.87 (0.79-0.96) | 91.3 | 0.013 | 0.79 |
| >18 | 15 | 0.92 (0.87-0.97) |  |  |  |
| <u>Median PFS (Month)*</u> |  |  |  |  |  |
| <=6 | 13 | 0.82 (0.74-0.91) | 90.6 | 0.017 | 0.85 |
| >6 | 10 | 0.99 (0.97-1.02) |  |  |  |
| <u>Median OS (Month)*</u> |  |  |  |  |  |
| <=12 | 8 | 0.78 (0.68-0.91) | 93.1 | 0.016 | 0.85 |
| >12 | 14 | 0.93 (0.89-0.98) |  |  |  |

1) Abbreviations: RR, relative risk; I<sup>2</sup>, percent residual variation due to heterogeneity; τ<sup>2</sup>, residual maximum likelihood estimates of between-study variance; CI, confidence interval; ECOG, Eastern Cooperative Oncology Group score; PFS, progression free survival; OS, overall survival.
2) \*Not all trials have the available information.
3) <sup>^</sup>0.64 is the median percentage.

**Figure 4.**
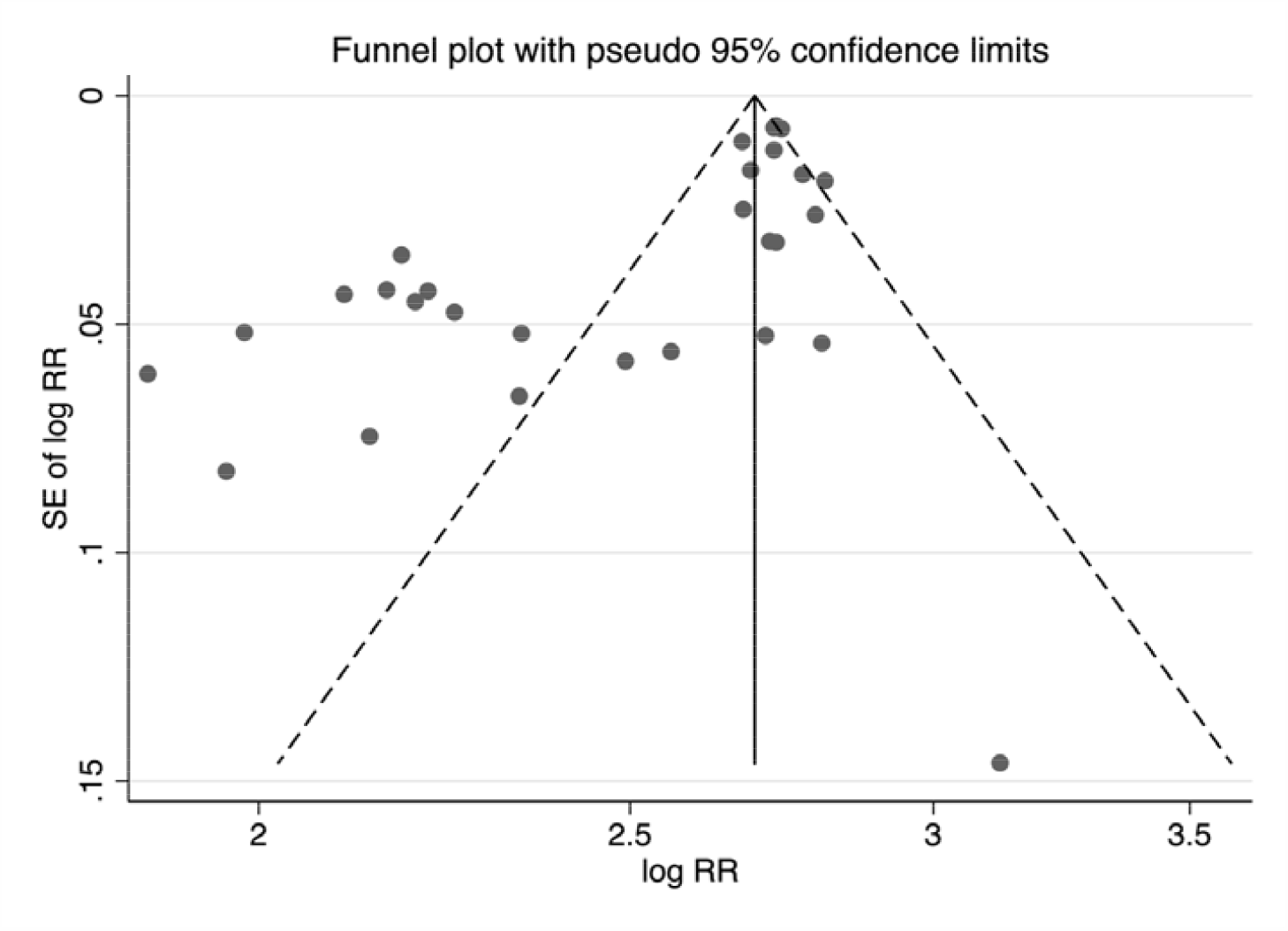
Funnel Plot for All Included 29 Studies Included in the Meta-Analysis.

## 4 Discussion

PD-1 and PD-L1 inhibitors are arguably the most important development in cancer therapy over the past decade. The indications for PD-1/PD-L1 inhibitors continue to expand across malignancies and disease settings, thus reshaping many of the previous standard-of-care approaches and bringing new hope to patients [94]. One of the costs of these advances is the emergence of a new spectrum of toxicities, which are often distinctly different from those of classical cytotoxic chemotherapy. They may involve any system of the body, and while they are generally manageable, they range from trivial to fatal [95]. A comprehensive understanding of the spectrum of toxicity of PD-1/PD-L1 inhibitors is critical. Awareness needs to be raised regarding the toxicity spectrum based on variables like toxicity grade, system and organ, treatment regimens in the intervention arm, treatment regimens in the control arm, drug type, cancer histotype, and drug dose and duration. Over the past few years, many clinical trials including several RCTs have investigated the therapeutic effects, safety and toxicities of PD-1/PD-L1 inhibitors. However, the reporting of resulting toxicity was not comprehensive and not consistent between different studies. The significance of our present meta-analysis lies in that it is a systematic integration and synthesis of existing related RCTs.

Our results suggest that PD-1/PD-L1 inhibitors are associated with a lower risk of gastrointestinal toxicity and hematologic toxicity, and a higher risk of respiratory toxicity and endocrine toxicity than chemotherapy and anti-CTLA-4 therapy. There have been several previous meta-analysis studies that have reported the toxicities of PD-1/PD-L1 inhibitors in recent years, however these have a series of limitations such as insufficient case number, uncontrolled single-arm study, inadequate sub-group analysis, etc. We used data from 29 randomized controlled trials that included a total of 16173 patients in this study. To the best of our knowledge, the present meta-analysis is the most comprehensive, detailed, and panoramic systematic review and meta-analysis of randomized controlled trials for toxicities of immune-checkpoint inhibitors.

For toxicities at any grade, gastrointestinal toxicity had the highest incidence proportion, followed by constitutional toxicity and then hematologic toxicity, which was probably driven by the anti-PD-1/PD-L1 plus chemotherapy trials. While for severe toxicities (Grade 3∼4), the hematologic toxicity ranked the highest. These differences were not unique to the intervention arm, reflecting the general tendency of every toxicity type and the influence of cytotoxic chemotherapy in the intervention arm (xx studies including anti-PD-1/PD-L1 plus cytotoxic chemotherapy). For most toxicity types based on organ system, the incidence proportions for patients in the intervention arm were lower than those in the control arm, which reinforces the general safety of PD-1/PD-L1 inhibitors compared with conventional chemotherapies. However, for respiratory toxicity, some kinds of cutaneous toxicity like pruritus and rash, arthralgia, and endocrine toxicity, patients in the intervention arm had significant higher incidence proportions than in the control arm. Of note, the comparative frequency of certain toxicities was not the same between any grade and Grade 3∼4. For example, when evaluating the rate of any grade of pyrexia, the incidence in the intervention arm was much higher than that in the control arm. However, for Grade 3∼4 pyrexia, the result was just the opposite, indicating that patients receiving PD-1/PD-L1 inhibitors were more likely to develop mild pyrexia than patients in the control arm, but less likely to develop severe cases.

Although most toxicities were broadly comparable among the PD-1/PD-L1 inhibitors, particular toxicity incidence changes varied with the drug types used in the intervention arm. From our results, PD-1 inhibitors did not increase or decrease the incidence of cough and colitis. However, cough incidence and colitis incidence both were increased significantly in patients who received a PD-L1 inhibitor. Rash incidence was increased in patients treated with pembrolizumab or PD-L1 Inhibitor, but had no significant changes for nivolumab (all grades: RR 1.36, 95% CI 0.99-1.88; Grade 3∼4: RR 0.51, 95% CI 0.25-1.04). Similarly, the incidence of any grade or Grade 3∼4 pneumonitis and hepatitis were increased in patients treated with pembrolizumab, rather than nivolumab and PD-L1 inhibitors. Our results were a breakthrough against Pillai et al.’s study published in 2017 [96]. They carried out a meta-analysis on studies of PD-1 and PD-L1 inhibitors (nivolumab and pembrolizumab, and atezolizumab, durvalumab and avelumab respectively) used as single agents in advanced lung cancer patients, published between 2000 and 2016. The toxicity profiles of the PD-1 and PD-L1 inhibitors appeared comparable, with overall toxicity incidences of 64% and 66%, respectively (P=0.8). However, 13% of patients undergoing treatment with PD-1 inhibitors and 21% of those given PD-L1 inhibitors developed various toxicities at Grades 3∼4 (P=0.15). The rate of toxicities resulting from the treatment of PD-1 inhibitors was 16%, and that seen with PD-L1 inhibitors was 11% (P=0.07), although the incidences of toxicities at Grades 3∼4 in the two groups were 3% and 5% respectively (P=0.4). Patients receiving PD-1 inhibitors experienced a higher incidence of pneumonitis than those who were given PD-L1 inhibitors (4% vs. 2%; P=0.01) [97].

Potential reasons for the differences of toxic effects between nivolumab and pembrolizumab might include the following. Firstly, although both nivolumab and pembrolizumab target epitopes on the PD-1 molecule and are of the IgG4 subclass, they do have some slight differences in their antibody molecular property. For nivolumab, binding is dominated by interactions with the PD-1 N-loop. Total buried surface 1487-1932.5 Å2; yet for pembrolizumab, binding dominated by interactions with PD-1 CD loop. Total buried surface 1774-2126 Å2. The epitope determines the drug’s molecular target and therefore its mode of action. The epitope difference between the two drugs may influence their on-target side effects. Secondly, the strength with which antibody binds target molecule alters the degree of on-target side effects. The two drugs’ affinities for recombinant human PD-1 protein (surface plasmon resonance) are different (nivolumab Kd=3.06 pM; pembrolizumab Kd=29 pmol/L). Thirdly, with antibody specificity decreases, off-target side effects become more likely. No binding to other members of superfamily were found for nivolumab, including CD28, ICOS, CTLA-4, and BTLA (ELISA); while the data was not available for pembrolizumab [98]. Fourthly, the apparent difference is not real and due to confounding factors of the individual trials, since this comparative toxicity analysis is based on a comparison of changes in toxicity incidence of trials involving a PD-1/PD-L1 inhibitor versus a control arm that most commonly was cytotoxic chemotherapy.

Between melanoma and lung cancer, most toxicity incidence differences between intervention arms and control arms were similar. Nevertheless, based on toxicity types, whether there were significant incidence changes was not always the same and sometimes the change direction could even be the opposite. For melanoma, patients in the intervention arm were at higher risks than those in the control arm to develop asthenia (all grades: RR 1.14, 95% CI 1.00-1.29); for lung cancer patients, however, they were at lower risks (all grades: RR 0.71, 95% CI 0.58-0.87). For melanoma, patients in the intervention arm were less likely to experience hepatotoxicity (Grade 3∼4: RR 0.85, 95% CI 0.53-1.36) and colitis (Grade 3∼4: RR 0.31, 95% CI 0.11-0.84); however, for lung cancer, they were more likely to develop these toxicities (hepatotoxicity: all grades RR 1.44, 95% CI 1.21-1.72; colitis: all grades RR 2.61, 95% CI 1.00-6.79, Grade 3∼4 RR 2.16, 95% CI 1.06-4.41). This phenomenon may be explained by at least three reasons: first, patients with different types of cancers have different homeostasis points and different tumor microenvironments, so their responses to drugs may be different. Second, the generation of different types of toxicities may involve different cellular or molecular targets and different signal transduction pathways, leading to discrepant toxic effects in patients with different types of cancers. Third, the control arm for lung cancer trials was typically cytotoxic chemotherapy but very often ipilimumab for melanoma.

Apart from the strength of this study that data were obtained from 29 formal RCTs published in internationally authoritative journals instead of other retrospective studies or single-arm studies to ensure the comparability of the included studies and avoid the risk of bias in this meta-analysis, the detailed subgroup analysis from a series of aspects was also a strength of this study. In addition to the above-mentioned stratification by system and organ, PD-1/PD-L1 inhibitor drug type, cancer histotype, and toxicity grade, we also performed a subgroup analysis stratified by the treatment regimen in the control arm. Among the included studies, the CTLA-4 inhibitor was used as the control in **6** studies. The CTLA-4 inhibitor is also an immunotherapy agent, but its target is different from PD-1/PD-L1 inhibitors, and there are also large differences in their downstream signal transduction pathways. According to Qin et al., the binding of CTLA-4 to B7 proteins competes CD28 costimulatory signals and eventually acts to impede excessive immunity; while PD-1 play critical roles in the maintenance of peripheral tolerance [99-101]. The engagement of PD-1 by its ligands results in the recruitment of Src homology 2 (SH2) domain containing phosphatases 1/2 (SHP1/2) and then inhibits T cell proliferation and cytokine secretion mediated by TCR [102]. Few studies compared the toxicities of these two kinds of immunotherapy. In 2017, Velasco et al. found that there were no significant differences in the RR between PD-1/PD-L1 inhibitors vs. CTLA-4 inhibitors for toxicities at both any grade and Grade 3∼4 except rash (PD-1/PD-L1 RR 1.59, 95% CI 0.90∼2.82; CTLA-4 RR 3.94 95% CI 3.02∼5.14; P=0.006) and colitis (PD-1/PD-L1 RR 2.47, 95% CI 0.90∼6.72; CTLA-4 RR 22.5 95% CI 6.37∼79.4; P=0.021) [103]. In our analysis, the differences between PD-1/PD-L1 inhibitors vs. CTLA-4 inhibitors were more detailed and accurate (see Results section 3.6), partly because our study included much more cases and we conducted a much more comprehensive subgroup analysis.

This meta-analysis harbored several limitations:

1. This is a meta-analysis at the study level; therefore, variables at the patient level were not included in the analysis. Thus, we could not establish risk factors associated with the development of toxicities. Also, the patients in studies selected for our meta-analysis were a select group of patients with good performance status who were recruited into clinical trials conducted at academic centers. The actual incidence of toxicities in patients with organ dysfunction and/or an impaired functional status is likely to be higher in clinical practice. Of course, “healthier” patients might have more robust immune systems and might actually have less autoimmune toxicity even though they may be less likely to have cytotoxicity from chemotherapy. However, the patient selection into the trails in our study is unlikely to introduce bias into the OR analysis of the toxicities.
2. Significant heterogeneity was observed in the included studies for some of the planned OR analyses. We minimized heterogeneity influence by using the random-effects model and also performed exploratory subgroup analyses based on type of toxicity grade, specific affected system and organ, treatment regimen, drug type, and cancer histotype. As there were differences in the risk of some individual AEs between the subgroups, the observed heterogeneity may be partially explained by the differences in these factors.
3. There was possible overlap in CTCAE definitions which prevents understanding the true rates of specific toxicities. For example, immune-related hepatitis could be captured as hepatitis or as an abnormal laboratory value (elevated aspartate transaminase and alanine transaminase) and immune-mediated colitis could be categorized as colitis or diarrhea. This could lead to potential uncertainty regarding the quality of the data, which will need to be addressed moving forward for studies of immune checkpoint inhibitors.
4. We assumed that the non-reporting of toxicity data was the result of either no events or non-measurement of the outcome. If a selective non-reporting mechanism were present, such that the toxicity data was measured but not reported based on the results, then there is the possibility that we may have overestimated the drugs’ safety [104].
5. We pooled data from studies that used different PD-1/PD-L1 inhibitors at variable doses so we may have missed differences in toxicity rates across drugs or based on dosage differences. Given the wide variation in drug and dose across studies, we were unable to perform subgroup analyses to examine these factors. However, we found little heterogeneity across studies for toxicity outcomes, suggesting little difference based on the specific drug or dose. Also, anti-PD-1/PD-L1 toxicity and efficacy were considered not dose dependent once above a level of 1 mg/kg.
6. Our study provides insight into the toxicities from treatment with PD-1 and PD-L1 inhibitors, which have revolutionized care for patients with cancer in the last few years. However, owing to the short interval of follow-up data currently available from clinical trials and a lack of clarity in the systematic capture of many toxicities, we are likely to have underestimated the true rates of toxicities. At the very least, we likely are underestimating toxicities from anti-PD-1/PD-L1 therapy that develop after prolonged treatment as well as those that occur after treatment discontinuation, since these tend not to be captured in the studies of efficacy at the time of trial result reporting. Moving forward, longer term follow-up and specific attention to a variety of immune-related toxicities may enhance our understanding.
7. Due to heterogeneities found in the meta analyses, multiple meta-regressions were conducted to investigate the possible source of heterogeneities. However, we only found that male patient percentage was a possible source of heterogeneity. Our study also revealed evidence of heterogeneities and/or publication bias by funnel plot. There may be heterogeneities from another source which this article did not investigate comprehensively.

## 5 Conclusion and Perspective

This meta-analysis, which used a series of subgroup analysis strategies, has defined the differences of risks of all common toxicities between PD-1/PD-L1 inhibitors and the control. The incidence differences of toxicities are associated with systems and organs, toxicity types, toxicity grades, drug types, treatment regimens in intervention arms or in control arms, and cancer histotypes. This panorama overview of the toxicities of PD-1 and PD-L1 inhibitors can be used as a reference by clinicians and oncologists and may guide clinical practice. Future research should focus on taking effective targeted measures to decrease the risks of different toxicities for different patient populations.

## Supplemental Files

**Table S1** Previously Published Meta-Analysis Articles with Research Characteristics and Limitation.

**Table S2** Statistical Results of the Meta-Analysis on the Comparison of Various Toxicities between Immunotherapy and the Control.

## Data Availability

All data are from the peer-reviewed publication.

